# Epigenetic timing effects on child developmental outcomes: A longitudinal meta-regression of findings from the Pregnancy And Childhood Epigenetics Consortium

**DOI:** 10.1101/2024.02.29.24303506

**Authors:** Alexander Neumann, Sara Sammallahti, Marta Cosin-Tomas, Sarah E Reese, Matthew Suderman, Silvia Alemany, Catarina Almqvist, Sandra Andrusaityte, Syed H Arshad, Marian J Bakermans-Kranenburg, Lawrence Beilin, Carrie Breton, Mariona Bustamante, Darina Czamara, Dana Dabelea, Celeste Eng, Brenda Eskenazi, Bernard F Fuemmeler, Frank D Gilliland, Regina Grazuleviciene, Siri E Håberg, Gunda Herberth, Nina Holland, Amy Hough, Donglei Hu, Karen Huen, Anke Hüls, Jianping Jin, Jordi Julvez, Berthold V Koletzko, Gerard H Koppelman, Inger Kull, Xueling Lu, Léa Maitre, Dan Mason, Erik Melén, Simon K Merid, Peter L Molloy, Trevor A Mori, Rosa H Mulder, Christian M Page, Rebecca C Richmond, Stefan Röder, Jason P Ross, Laura Schellhas, Sylvain Sebert, Dean Sheppard, Harold Snieder, Anne P Starling, Dan J Stein, Gwen Tindula, Marinus H van IJzendoorn, Judith Vonk, Esther Walton, Jonathan Witonsky, Cheng-Jian Xu, Ivana V Yang, Paul D Yousefi, Heather J Zar, Ana C Zenclussen, Hongmei Zhang, Henning Tiemeier, Stephanie J London, Janine F Felix, Charlotte Cecil

## Abstract

DNA methylation (DNAm) is a developmentally dynamic epigenetic process, yet we still know little about how epigenetic effects on health outcomes vary over time; whether DNAm alterations during certain periods of development are more informative than others; and whether epigenetic timing effects differ by outcome. To address these questions, we applied longitudinal meta-regression to published meta-analyses from the PACE consortium that examine DNAm at multiple time points (prospectively at birth and cross-sectionally in childhood) in relation to the same child outcome (ADHD, general psychopathology, sleep, BMI, asthma). Our findings reveal three new insights: (i) across outcomes, effects sizes are larger when DNAm is measured in childhood compared to at birth; (ii) higher effect sizes do not necessarily translate into more significant findings, as associations also become noisier in childhood for most outcomes (i.e. showing larger standard errors); and (iii) DNAm signals are highly time-specific while showing pleiotropy across health outcomes.

## Introduction

DNA methylation (DNAm) is an important epigenetic regulator of development and health. DNAm is influenced by both genetic^1,2^ and environmental factors, beginning in utero (e.g. maternal smoking,^3^ stressful life events^4^, air pollution,^5^ or physical activity^6^). Alterations in DNAm have also been linked to a wide range of health outcomes across childhood, including asthma^7^, attention-deficit/hyperactivity disorder (ADHD)^8^, and body mass index (BMI)^9^. Together, these properties make DNAm an attractive biological process in the search for both biomarkers and mediators of disease risk.

DNAm is highly dynamic during development – a property that makes it particularly interesting, but also challenging to study. Mulder et al.^10^ estimated that over half of DNAm sites show changes in methylation from birth to 18 years of age, often following a non-linear trajectory. Furthermore, in around a third of DNAm sites, the degree of change varies between individuals, perhaps reflecting exposure to different postnatal environments, genetic variation or stochastic processes.^11^ Yet, most observational studies linking DNAm to health phenotypes measure DNAm only once.^12^ Thus, it is largely unknown (i) whether the relationship between DNAm and health outcomes varies across development (ii) at which developmental periods DNAm profiles could be most informative for a given health outcome, and (iii) to what extent DNAm-health associations at one time point can be generalized to other time points.

Population-based cohorts have emerged as a powerful tool for the study of DNAm-health associations, due to their relatively large sample sizes and longitudinal follow-up. In most pediatric population studies, DNAm is either measured in cord blood samples at birth and associated with a child outcome at a later time point (i.e. prospective epigenome-wide association study [EWAS]) *or* DNAm is measured from a blood sample at the same time point as the child outcome (i.e. cross-sectional EWAS). Theoretical arguments exist for either design. On the one hand, DNAm measured in cord blood at birth coincides with a developmentally sensitive period and may reflect causal effects of genetic and *in utero* environmental factors that can influence risk of later outcomes.^13^ Furthermore, reverse causation scenarios are less likely, given that outcomes in childhood are unlikely to affect methylation profiles at birth. However, cross-sectional EWASs during childhood may result in a stronger association signal, due to the temporal proximity between predictor and outcome, a larger accumulation of environmental effects (prenatal and postnatal), or the potential for DNAm patterns in childhood to reflect both a cause and consequence of poor health (reverse causality). Cord and peripheral blood also represent different tissues, with different cell compositions (e.g., nucleated red blood cells being present in cord, but not peripheral blood), which may contribute to differences in associations of DNAm with health outcomes.^14^ However, it is challenging to fully separate the influence of tissue versus timing, as for example cord blood is only available at birth, and early cell-type changes are in part developmentally regulated.^14,15^

Recently, the Pregnancy And Childhood Epigenetics (PACE) Consortium^16^ published five multi-cohort EWAS meta-analyses that investigated DNAm using *both designs* in relation to the same child outcome, spanning mental and physical health domains, namely: ADHD,^8^ general psychopathology (measured as a latent factor; GPF),^17^ sleep duration,^18^ body mass index (BMI)^9^ and asthma^7^. Results from these previous studies can be summarized as follows (Table 1): for ADHD, there were more hits for DNAm at birth rather than in childhood (i.e prospective EWAS showed more hits than cross-sectional EWAS); whereas the opposite was true for BMI and asthma (i.e. prospective EWAS showed fewer hits than cross-sectional EWAS). For GPF and sleep, results were mostly null at either time point. Together, these findings point to the potential existence of epigenetic ‘timing effects’ on child health.

**Table 1.** Published epigenome-wide association studies of child developmental outcomes from PACE, jointly re-analyzed in present study.

| Study | Outcome | Age | Instrument | Methylation | Model | Covariates | Meta-analysis | Sig. Threshold | Birth EWAS |  | Childhood EWAS |  |
| --- | --- | --- | --- | --- | --- | --- | --- | --- | --- | --- | --- | --- |
|  |  |  |  |  |  |  |  |  | n | n<sub>cpg</sub> | n | n<sub>cpg</sub> |
| Neumann et al. (2020) | ADHD | 5-15 | Paternal questionnaire | Beta | LMM, LM | sex, gest. age, age*, mat. age, mat. edu., mat. smoking, cell prop., batch | HE-RE | Bonf. | 2477 | 9 | 2374 | 0 |
| Rijlaarsdam et al. (2022) | GPF | 6-12 | Paternal questionnaire | Beta (Trimmed) | RLM | sex, gest. age, age, mat. age, mat. edu., mat. smoking, cell prop., ancestry, batch | FE | Bonf. | 2178 | 0 | 2190 | 1 |
| Sammallahti et al. (2022) | Sleep | 4-13 | Paternal questionnaire | Beta (Trimmed) | LM | sex, age, mat. age, mat. edu., mat. smoking, cell prop., ancestry, batch | FE | FDR | 3658 | 0 | 2539 | 0 |
| Vehmeijer et al. (2020) | BMI | 2-10 | Measured | Beta | RLM | sex<sup>#</sup>, gest age, age<sup>#</sup>, mat. age, mat. edu., mat. smoking, mat. BMI, parity, birth weight, breastfeeding, cell prop., ancestry, batch | FE | Bonf. (FDR) | 4641 | 1 (1) | 3406 | 1 (10) |
| Reese et al. (2019) | Asthma | 5-17 | Diagnosis | Beta | Logistic | sex, mat. age, mat. edu., mat. smoking, cell prop., batch | FE | FDR | 3572 | 9 | 2834 | 179 |

\* Covariate only used in school-age analyses

Despite these intriguing findings, the studies’ main goal was to maximize identification of health-relevant DNAm sites at each time point, rather than systematically quantify temporal changes of DNAm-health associations. Addressing this aim would require specific analyses that were not originally performed, including quantitatively comparing effect sizes between time points, accounting for sample size imbalances that affect statistical power per time point, and examining potential statistical and biological factors contributing to temporal differences in DNAm-health associations. Furthermore, no comparison has been performed *across* studies, to establish how temporal patterns may vary for different child health outcomes, and whether methylation signals for one outcome correlate with that for other outcomes (i.e., indicating pleiotropy/shared epigenetic effects).

Here, we re-analyzed the five PACE meta-analyses on ADHD, GPF, sleep, BMI and asthma (N_pooled_=2178-4641, 26 cohorts) to explore timing effects on DNAm-health associations during development. For each outcome, we integrated results from the prospective EWAS (cord blood DNAm at birth) and the cross-sectional EWAS (whole blood DNAm in childhood) into a longitudinal meta-regression model. This model enabled us to systematically quantify changes in effect sizes and statistical significance between time points, and also explore a range of factors that may contribute to the observed temporal trends. We then performed correlation analyses to estimate the consistency of DNAm associations *between time points* (i.e. in order to assess generalizability of epigenetic signals from one time point to another) and *across child health outcomes* (i.e. to explore presence of shared DNAm associations).

## Results

### How do EWAS effect sizes change from birth to childhood?

We applied multilevel meta-regression models in which regression coefficients (β) from the prospective and cross-sectional EWAS were pooled across cohorts and regressed on a variable indicating whether the estimate pertains to birth or childhood DNAm. This model therefore quantified the DNAm associations at birth, in childhood, as well as the differences in associations between time points. β here represents the difference in child health outcomes in standard deviations (SD) between no to full methylation in the case of continuous variables or as odds ratio for the categorical outcome asthma. We focused on effect sizes as defined by the absolute regression coefficient at birth |β_birth_| or in childhood |β_childhood_|. Furthermore, we characterized global trends defined by mean statistics, averaged across all autosomal DNAm sites tested. Tables S1 and S2 show an overview of included cohorts and the overlap between time points and outcomes.

For DNAm at birth, mean effect sizes across DNAm sites ranged from 0.77 (BMI) to 1.23 (GPF) for continuous measures (Table 2; Figures 1,2,S1,S2). Averaged across phenotypes, 10% higher methylation was associated with a 0.10SD outcome difference. For asthma, mean log(odds) were 2.70, which corresponds to a 10% methylation difference being associated with 1.30 lower/higher odds of receiving an asthma diagnosis.

**Figure 1.**
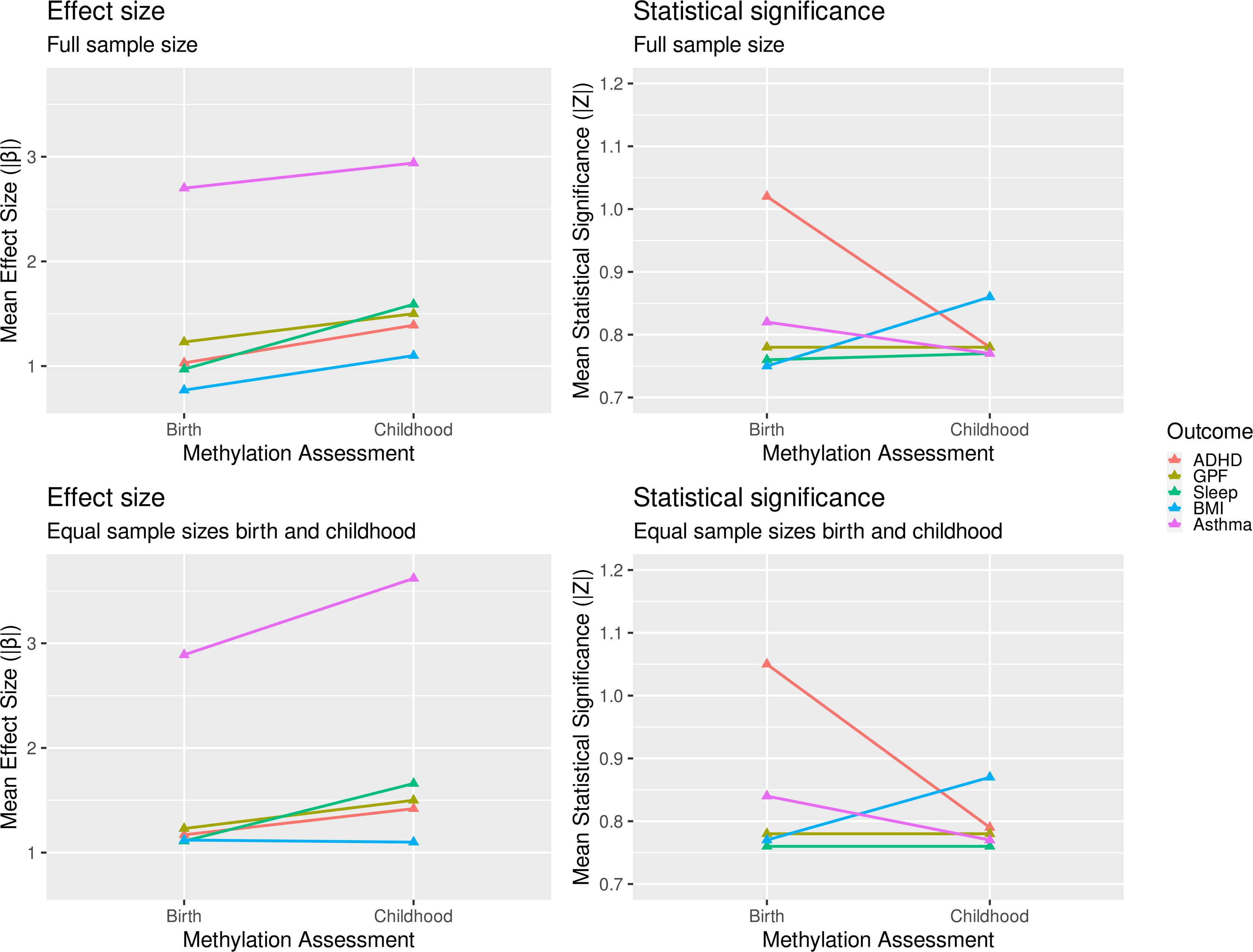
Mean effect sizes and statistical significance for DNAm at birth and in childhood. Mean effect sizes (left column) and mean statistical significance (right column) across all tested autosomal DNAm sites per outcome (color) and time point. Upper row displays results from analyses utilizing maximum available sample sizes. Lower row displays results from analyses with cohorts removed to achieve equal sample sizes at both time points. Effect size is given as absolute regression coefficient (|β̄|), representing the difference in child health outcomes in SD between full or no methylation in the case of continuous outcomes (ADHD, general psychopathology, sleep duration and BMI), or log(odds ratio) for categorical outcomes (asthma diagnosis). Statistical significance is given as mean absolute Z-values.

**Figure 2.**
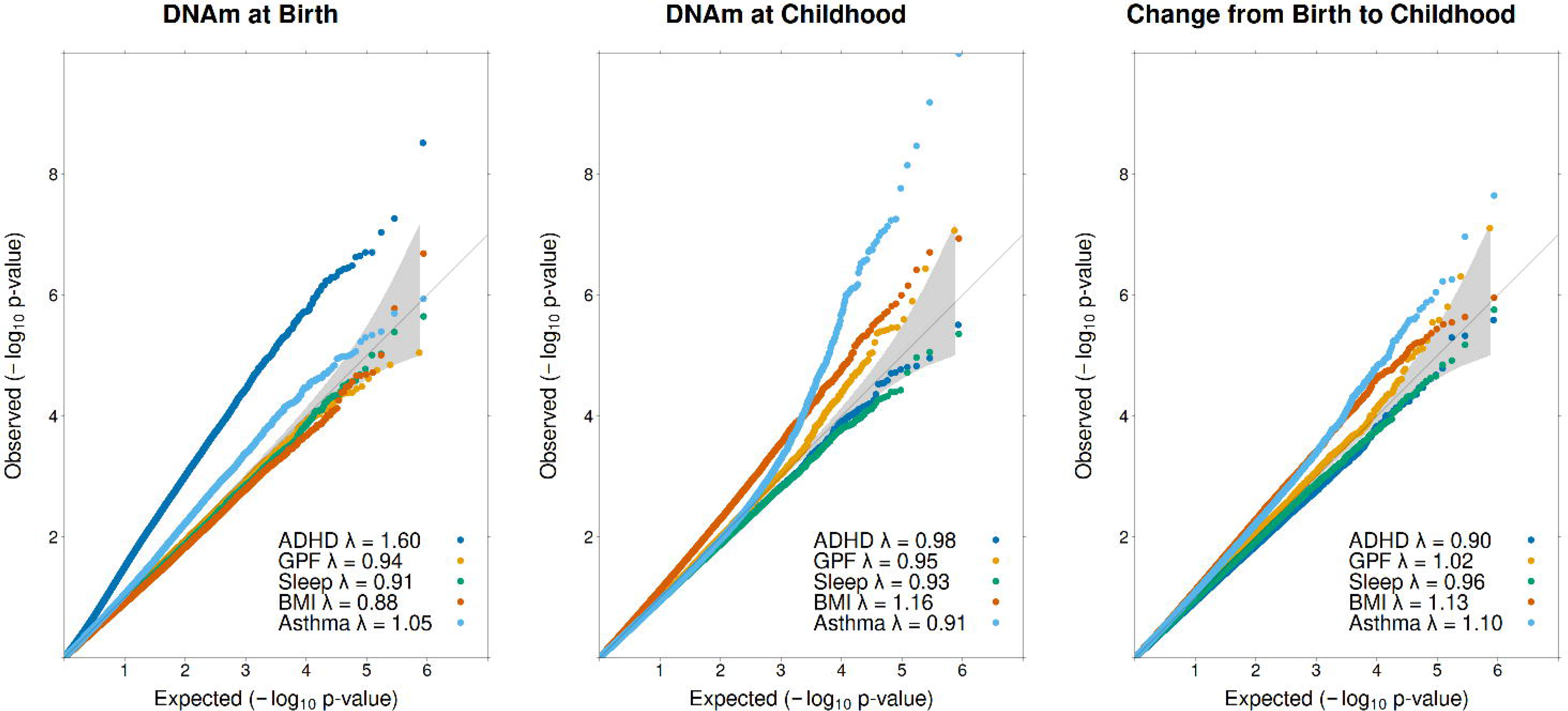
QQ-plots. Distribution of observed p-values (y-axis) vs expected (x-axis). Diagonal represents expected distribution of p-values by chance. Upwards deviations indicate higher presence of lower p-values than expected assuming a null effect. Distributions are given for DNAm effects at birth (left), in childhood (middle) and for change in effect between birth and childhood (right) per outcome (color). Grey are displays the 95% confidence interval of the null distribution.

**Table 2.** Association between DNA methylation either at birth or in childhood and child developmental outcomes (full sample)

| Outcome | DNAm at Birth (Prospective EWAS) |  |  |  |  |  |  | DNAm in Childhood (Cross-sectional EWAS) |  |  |  |  |  | Change between time points |  |  |  |  |
| --- | --- | --- | --- | --- | --- | --- | --- | --- | --- | --- | --- | --- | --- | --- | --- | --- | --- | --- |
|  | n<sub>cpg</sub> | n | n<sub>cohorts</sub> | mean | mean | mean | n<sub>cpg</sub> p<0.05<br>(FDR/bonf.) | n | n<sub>cohorts</sub> | mean | mean | mean | n<sub>cpg</sub> p<0.05<br>(FDR/bonf.) | n<sub>cohorts</sub><br>both | Δβ | N<sub>cpg</sub><br>+Δβ | N<sub>cpg</sub><br>-Δβ | Δz |
|  |  |  |  | β<br>(abs.) | SE | z |  |  |  | β<br>(abs.) | SE | z |  |  |  |  |  |  |
| ADHD | 430327 | 2477 | 6 | 1.03 | 1.10 | 1.02 | 57339<br>(896/3) | 2374 | 5 | 1.39 | 1.76 | 0.78 | 19034<br>(0/0) | 3 | 0.36 | 10542(0/0) | 6841(0/0) | -0.23 |
| GPF | 372292 | 2178 | 4 | 1.23 | 1.59 | 0.78 | 16549<br>(0/0) | 2190 | 5 | 1.50 | 1.98 | 0.78 | 17767<br>(1/1) | 3 | 0.27 | 13375(1/1) | 6475(0/0) | 0.01 |
| Sleep | 431159 | 3658 | 10 | 0.97 | 1.30 | 0.76 | 17399<br>(0/0) | 2539 | 5 | 1.59 | 2.06 | 0.77 | 18113<br>(0/0) | 4 | 0.63 | 14447<br>(0/0) | 5171(0/0) | 0.01 |
| BMI | 435652 | 4102 | 14 | 0.77 | 1.04 | 0.75 | 16012<br>(0/0) | 3406 | 11 | 1.10 | 1.29 | 0.86 | 30615<br>(2/1) | 6 | 0.33 | 23793(0/0) | 6634(0/0) | 0.11 |
| Asthma | 432728 | 3065<br>(631) | 7 | 2.70 | 3.44 | 0.82 | 26112<br>(0/0) | 2834<br>(631) | 9 | 2.94 | 3.92 | 0.77 | 18605<br>(66/11) | 0 | 0.24 | 18024(2/2) | 9678(2/0) | -0.06 |

**ADHD** Attention Deficit Hyperactivity Disorder
**GPF** General Psychopathology Factor
**BMI** Body Mass Index
**n<sub>cpg</sub>** Number of CpG sites tested
**n** Sample size (cases)
**n<sub>cohorts</sub>** Number of cohorts
**mean $\beta$ (abs.)** The mean absolute regression coefficient across DNAm sites. $\beta$ represents the expected difference in the outcome in SD when CpG sites is fully methylated compared to no methylation. For asthma, $\beta$ represents the log(odds) difference
**Mean SE** Mean Standard Error
**Mean z** Mean z values across CpG sites. $z=\beta/SE$ indicating statistical significance

Compared to DNAm at birth, mean effect sizes for DNAm in childhood were consistently *higher* across all tested outcomes (Table 2,3; Figure 1,2,S1,S2), ranging from 1.10 (BMI) to 1.59 (Sleep) for continuous outcomes and an log(odds) of 2.94 (odds ratio of 1.34) for asthma. When quantifying this *difference* in effect sizes between birth and childhood, the smallest mean difference was observed for BMI (|β̅_childhood_|=1.10 vs |β̅_birth_|=0.77) and the largest difference for sleep (|β̅_childhood_|=1.59 vs |β̅_birth_|=0.97). Aggregating across continuous outcomes, mean effect sizes were 40% higher in childhood, with an outcome difference of 0.14SD per 10% methylation. For asthma, the odds ratio increased from 1.30 to 1.34. Table S3 shows effect size comparisons across percentiles.

**Table 3.** Comparison of birth EWAS (i.e. prospective analysis) versus childhood EWAS (i.e. cross-sectional analysis): Overview of study findings.

| Outcome | Change from birth to childhood EWAS |  | Potential contributing factors |  | Sensitivity analyses |  | Correlation analyses |  |
| --- | --- | --- | --- | --- | --- | --- | --- | --- |
|  | Effect size | Statistical significance | EWAS sample size | Between-study heterogeneity | Do results hold when making N equal across time points? | Do results hold within single longitudinal cohort (ALSPAC) | Correlations between time points (i.e. stability) | Correlations with other phenotypes |
| ADHD | ↑ | ↑ | = | ↑ | ? | ? | ? ($r_s = 0.31$) | ? ($r_s = -0.18-0.35$) |
| GPF | ↑ | = | = | = | ? | ? | ≠ ($r_s = 0.08$) | ? ($r_s = -0.07-0.35$) |
| Sleep | ↑ | = | ↓ | ↑ | ? | ? | ≠ ($r_s = 0.06$) | ≠ ($r_s = -0.04-0.06$) |
| BMI | ↑ | ↑ | ↓ | ↑ | ? | ? | ≠ ($r_s = 0.05$) | ? ($r_s = -0.18-0.06$) |
| Asthma | ↑ | ↑ | = | ↑ | ? | ? | ≠ ($r_s = -0.04$) | ? ($r_s = -0.16-0.21$) |

While these effect size figures provide a global view of genome-wide association change, they do not take into account statistical precision (i.e., standard error (SE)). Another way to quantify DNAm differences at birth versus in childhood is by counting the number of sites at which DNAm effect sizes increase or decrease over time based on a chosen p-value threshold of change. Among probes that showed at least a nominally significant difference between time points, there were 1.5-3x more DNAm sites with a larger as opposed to smaller effect size in childhood across health outcomes. (Table 2, Figures 2,S1-S4). To test the robustness of this approach, we also examined the ratio of DNAm sites that show an effect size increase vs decreases over time across different change p-value thresholds from no thresholding to p<0.0001 (Figure S4). We observed that the ratio is always positive (i.e. more DNAm showing an increase in effect size over time) – a trend that becomes stronger as the threshold becomes more stringent (lower p-values).

For the DNAm sites that showed at least nominally significant change over time, we also examined the direction of association with the health outcome, and whether this direction was consistent or not across time points. The most common pattern was a null or small effect at birth, followed by a positive association in childhood (Table S3). This applied to all outcomes, except BMI. Here the most frequent pattern was a switch from a positive association at birth to a negative association in childhood.

Three DNAm sites showed a genome-wide significant change in association. Cg11945228 in *BRD2* had no association with GPF at birth (β_birth_=5.28, SE=3.76, p=0.16), but became genome-wide significantly associated in childhood (β_childhood_=-37.00, SE=6.91, p=8.58*10^-8^), a significant change (p=7.68*10^-8^). Similarly, cg10644885 in *ACP5* had a significant change (p=2.25*10^-8^) from no association with asthma at birth (β_birth_=-0.56, SE=1.19, p=0.64) to genome-wide significance in childhood (β_childhood=_-15.00, SE=2.29, p=5.57*10^-11^). In addition, cg22708087 in *FRY* changed from a positive association with asthma at birth (β_birth_=7.47, SE=1.80, p=3.42*10^-5^) to a negative association in childhood (β_childhood_=-12.64, SE=3.32, p=1.44*10^-4^). This change was genome-wide significant (p=1.06*10^-7^). For all three genome-wide significantly changing DNAm sites, absolute effect sizes were larger in childhood.

### Do changes in effect size correspond with changes in the ability to identify significant associations?

While mean effect sizes were robustly larger for DNAm in childhood compared to DNAm at birth for all outcomes, this did not necessarily translate into more significant associations, as quantified by higher Z test-statistics (equal to lower p-values) (see also Tables 2,3 and Figures 1,2,S1,S2).

#### ADHD

DNAm at birth showed the strongest association signal with ADHD, as evidenced by a mean Z-value of 1.02 and the identification of the largest number of significant associations at all tested thresholds (Bonferroni, FDR, nominal). Despite an increase in effect sizes from birth to childhood, the mean Z-value dropped (1.02 at birth vs 0.78 in childhood). No CpG site was identified as genome-wide significant in the cross-sectional EWAS and the number of nominally significant sites was 3-fold lower (n_cpg-birth_ =57,339 vs n_cpg-childhood_=19,034).

#### GPF

The mean Z-value remained constant at 0.78 for both time points, and the number of nominally significant sites remained similar. No DNAm site reached genome-wide significance at birth, and one DNAm site reached genome-wide significance when assessed in childhood.

#### Sleep

Mean Z-values for sleep did not differ between time-points and the number of nominally significant sites remained similar, with no genome-wide significant hits at either time point.

#### BMI

For BMI the higher DNAm effect sizes in childhood corresponded with a higher statistical significance. This is also reflected by the doubling of nominally significant associations from birth to childhood (15,978 to 30,615), as well as by the presence of genome-wide hits in childhood, but not at birth.

#### Asthma

The mean Z-values and number of nominally significant sites were somewhat larger at birth than childhood. While this reflects the genome-wide trend, it is important to emphasize that the number of probes with genome-wide significance was much larger for DNAm in childhood (66 hits in the cross-sectional EWAS vs 0 hits in the prospective EWAS).

#### What explains these outcome-specific patterns?

We searched potential explanations for why statistical significance did not necessarily increase over time, or even decreased for specific outcomes, despite effect size increases. Z- and therefore p-values represent the ratio between effect size and statistical uncertainty. We found that standard errors (SE) increased from birth to childhood either to a disproportionately larger (ADHD, asthma) or similar (GPF, sleep) extent as the effect size increased (Tables 2,3), i.e. only for BMI did the increase in effect size outpace the increase in SE leading on average to more statistical significance.

Next, we investigated potential sources for the SE increase. The first was s*ample size*, which was unequal between EWAS time points for some outcomes. For GPF, total sample size was very similar, and for asthma the number of cases was also equal between time points. However, especially for sleep and BMI, sample sizes were much lower for DNAm measured in childhood, which increases SE. In sensitivity analyses we removed cohorts (Table S1) to achieve equal sample sizes between time points. Interestingly, patterns remained largely the same, i.e., with only BMI showing corresponding increases in both effect sizes and statistical significance over time (Table S4).

Second, we examined *between-study heterogeneity*, which tends to increase SE. We fit random slope models, allowing for different amounts of heterogeneity at different DNAm assessment periods. Indeed, we generally observed an increase in between-study heterogeneity for all outcomes over time, except for GPF (Table S5), suggesting that it may partly influence differences in EWAS signal between time points. We examined this possibility by re-computing meta-regression analyses using a single cohort. We chose ALSPAC, as it was the largest cohort contributing to all analyses with similar sample sizes at birth and childhood. Overall, the pattern of results in ALSPAC corresponded to the meta-analytical results for all outcomes, suggesting that observed temporal differences are unlikely to be solely explained by EWAS cohort composition in the meta-analyses.

### How do epigenetic signals correlate across time points and child outcomes?

To test the consistency of epigenetic associations over time and across outcomes, we computed spearman correlations (rs) between the regression coefficients of all time points and outcomes (Figure 3). For ADHD, estimates at birth correlated modestly with those in childhood (rs=0.31). For all other outcomes, estimates between time points were uncorrelated (rs<0.08). The coefficients in the ADHD analysis correlated most with the coefficients for other outcomes. For instance, the EWAS signal at birth for ADHD was positively correlated with the signal at birth for GPF (rs=0.35) and asthma (rs=0.21), but negatively correlated with the EWAS signal in childhood of BMI (rs=-0.18) and asthma (rs=-0.16).

**Figure 3.**
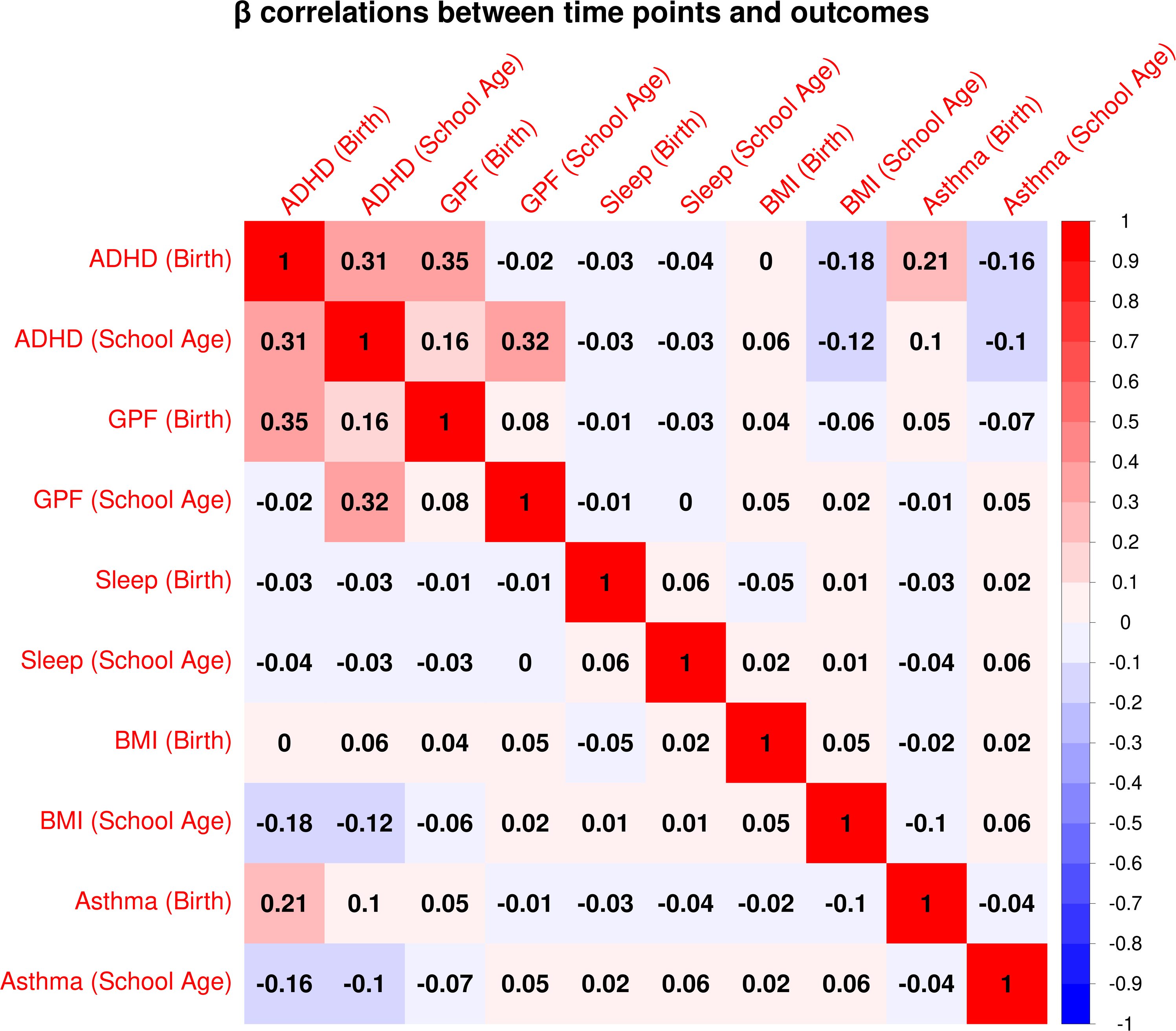
Correlations between DNAm effects at birth and childhood and across outcomes. This correlation matrix displays spearman correlations between regression coefficients for DNAm at birth and childhood and across outcomes. Intensity of red represents higher positive correlations and blue lower negative correlations.

Overlap between cohorts contributing to analyses at the same time point tended to be larger than between time points (Table S2). This may have led to an underestimation of correlations between time points. To test this, we re-ran correlation analyses within ALSPAC (as the largest cohort with repeated DNAm measures), and found that between time-point correlations remained low for GPF, sleep and BMI (rs<0.12) and modest for ADHD (rs=0.25) (Figure S5). Asthma could not be tested, due to unavailable analyses in childhood.

### Which biological pathways are involved in health-related DNAm patterns that change from birth to childhood?

We performed gene ontology enrichment analyses to probe the potential biological relevance of temporal changes in DNAm-health associations and to examine the possibility that we may be mainly picking up tissue differences (as opposed to developmental/ temporal differences) between birth and childhood. We selected sites that (i) were nominally associated with an outcome at either time point and (ii) showed at least nominally significant change in associations from birth to childhood. Notably for ADHD, GPF and sleep, neural features stand out among the top 10 pathways (e.g. cerebral cortex and neuron development, enrichment for synapses and dendrites, Table 4). While neural pathways also rank highly for BMI and asthma, other more general cell processes such as morphogenesis are prominently represented. However, no pathway was significant after adjustment for multiple testing of all 22,560 GO terms. See Tables S6-S11 for all pathways with nominal significance.

**Table 4.**
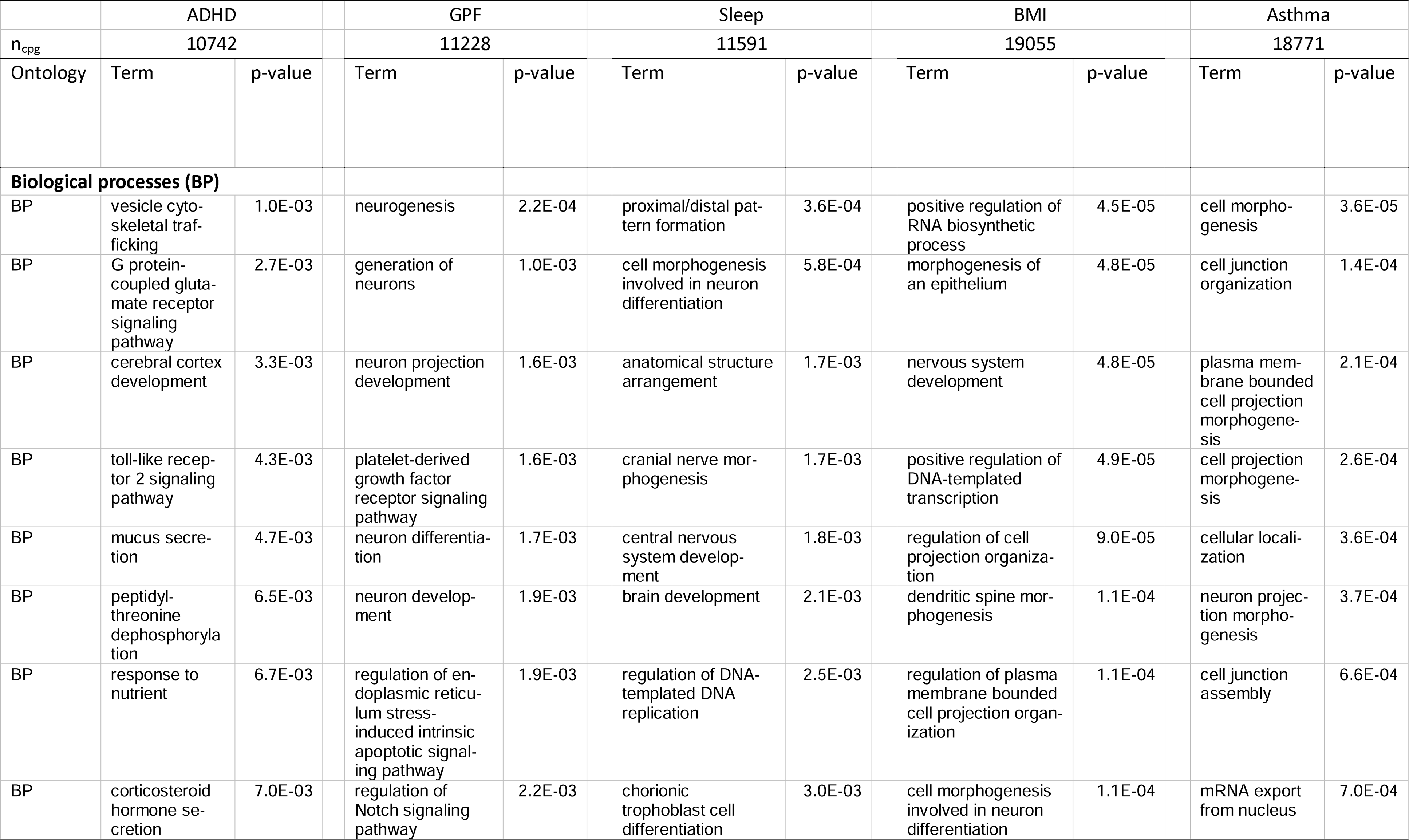
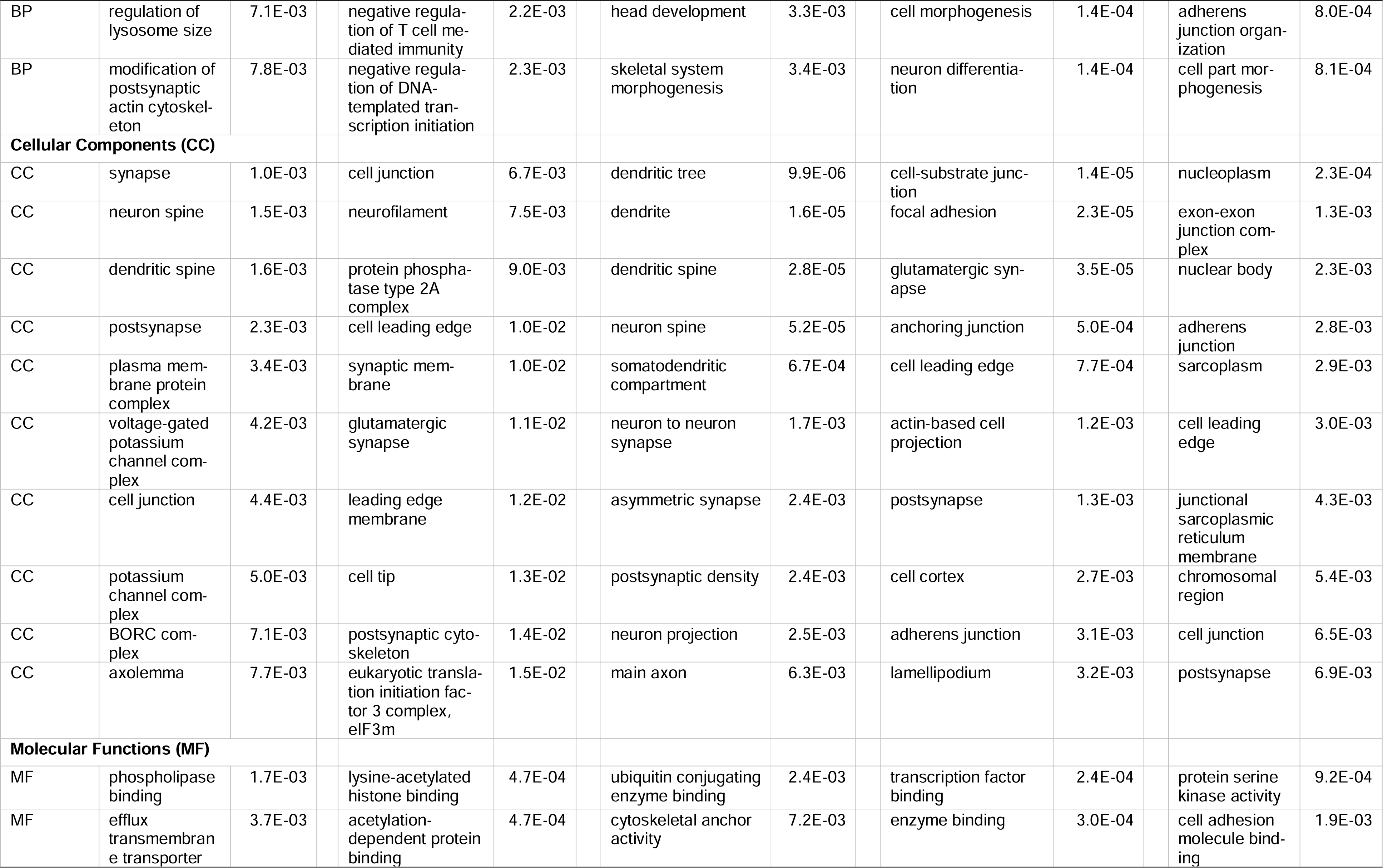
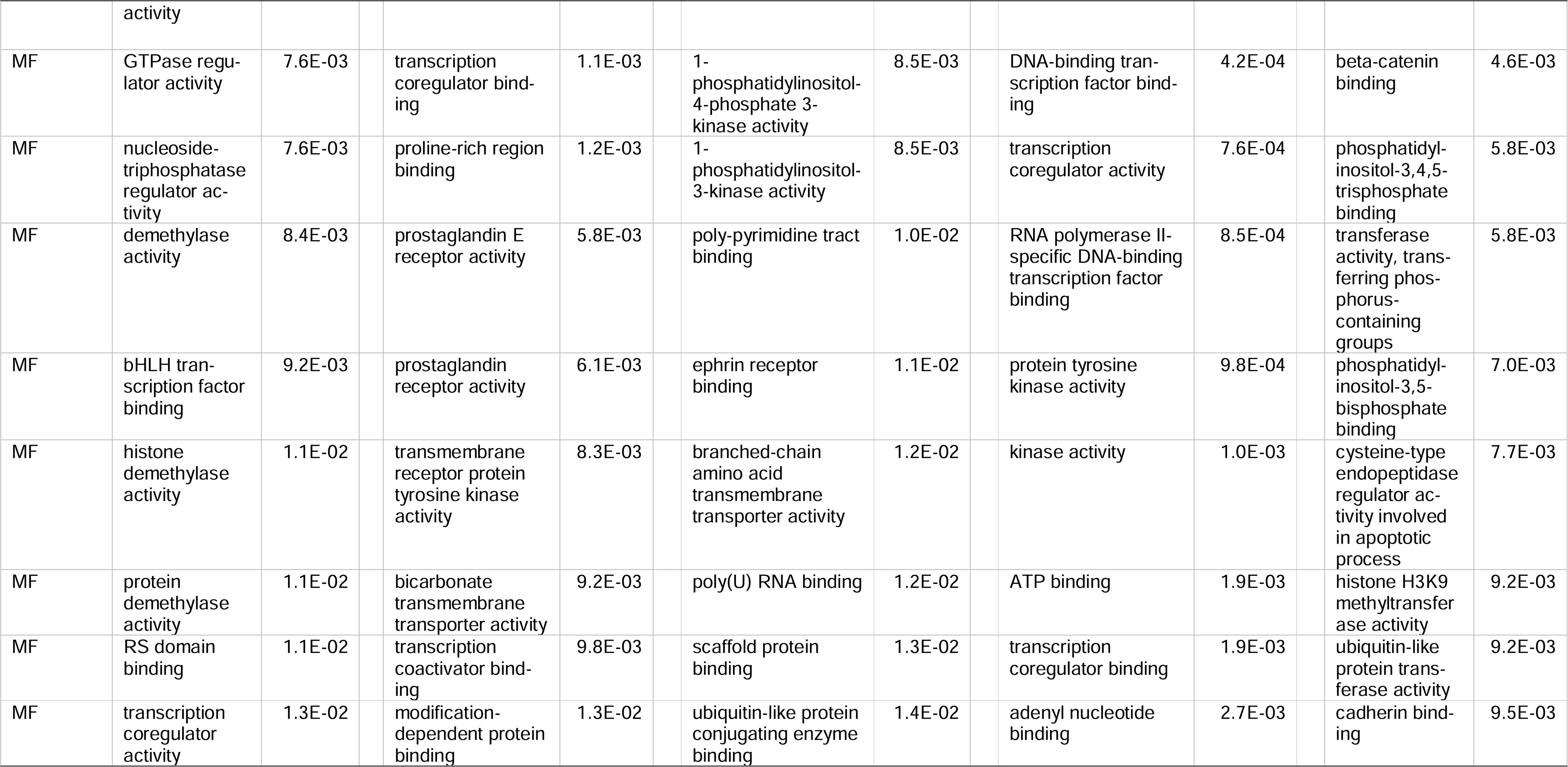
Gene ontology enrichment analyses: top 10 terms for phenotype-associated DNAm sites showing change from birth to childhood.

|  | ADHD |  | GPF |  | Sleep |  | BMI |  | Asthma |  |
| --- | --- | --- | --- | --- | --- | --- | --- | --- | --- | --- |
| n<sub>cpg</sub> | 10742 |  | 11228 |  | 11591 |  | 19055 |  | 18771 |  |
| Ontology | Term | p-value | Term | p-value | Term | p-value | Term | p-value | Term | p-value |
| **Biological processes (BP)** |  |  |  |  |  |  |  |  |  |  |
| BP | vesicle cytoskeletal trafficking | 1.0E-03 | neurogenesis | 2.2E-04 | proximal/distal pattern formation | 3.6E-04 | positive regulation of RNA biosynthetic process | 4.5E-05 | cell morphogenesis | 3.6E-05 |
| BP | G protein-coupled glutamate receptor signaling pathway | 2.7E-03 | generation of neurons | 1.0E-03 | cell morphogenesis involved in neuron differentiation | 5.8E-04 | morphogenesis of an epithelium | 4.8E-05 | cell junction organization | 1.4E-04 |
| BP | cerebral cortex development | 3.3E-03 | neuron projection development | 1.6E-03 | anatomical structure arrangement | 1.7E-03 | nervous system development | 4.8E-05 | plasma membrane bounded cell projection morphogenesis | 2.1E-04 |
| BP | toll-like receptor 2 signaling pathway | 4.3E-03 | platelet-derived growth factor receptor signaling pathway | 1.6E-03 | cranial nerve morphogenesis | 1.7E-03 | positive regulation of DNA-templated transcription | 4.9E-05 | cell projection morphogenesis | 2.6E-04 |
| BP | mucus secretion | 4.7E-03 | neuron differentiation | 1.7E-03 | central nervous system development | 1.8E-03 | regulation of cell projection organization | 9.0E-05 | cellular localization | 3.6E-04 |
| BP | peptidyl-threonine dephosphorylation | 6.5E-03 | neuron development | 1.9E-03 | brain development | 2.1E-03 | dendritic spine morphogenesis | 1.1E-04 | neuron projection morphogenesis | 3.7E-04 |
| BP | response to nutrient | 6.7E-03 | regulation of endoplasmic reticulum stress-induced intrinsic apoptotic signaling pathway | 1.9E-03 | regulation of DNA-templated DNA replication | 2.5E-03 | regulation of plasma membrane bounded cell projection organization | 1.1E-04 | cell junction assembly | 6.6E-04 |
| BP | corticosteroid hormone secretion | 7.0E-03 | regulation of Notch signaling pathway | 2.2E-03 | chorionic trophoblast cell differentiation | 3.0E-03 | cell morphogenesis involved in neuron differentiation | 1.1E-04 | mRNA export from nucleus | 7.0E-04 |
| BP | regulation of lysosome size | 7.1E-03 | negative regulation of T cell mediated immunity | 2.2E-03 | head development | 3.3E-03 | cell morphogenesis | 1.4E-04 | adherens junction organization | 8.0E-04 |
| BP | modification of postsynaptic actin cytoskeleton | 7.8E-03 | negative regulation of DNA-templated transcription initiation | 2.3E-03 | skeletal system morphogenesis | 3.4E-03 | neuron differentiation | 1.4E-04 | cell part morphogenesis | 8.1E-04 |
| **Cellular Components (CC)** |  |  |  |  |  |  |  |  |  |  |
| CC | synapse | 1.0E-03 | cell junction | 6.7E-03 | dendritic tree | 9.9E-06 | cell-substrate junction | 1.4E-05 | nucleoplasm | 2.3E-04 |
| CC | neuron spine | 1.5E-03 | neurofilament | 7.5E-03 | dendrite | 1.6E-05 | focal adhesion | 2.3E-05 | exon-exon junction complex | 1.3E-03 |
| CC | dendritic spine | 1.6E-03 | protein phosphatase type 2A complex | 9.0E-03 | dendritic spine | 2.8E-05 | glutamatergic synapse | 3.5E-05 | nuclear body | 2.3E-03 |
| CC | postsynapse | 2.3E-03 | cell leading edge | 1.0E-02 | neuron spine | 5.2E-05 | anchoring junction | 5.0E-04 | adherens junction | 2.8E-03 |
| CC | plasma membrane protein complex | 3.4E-03 | synaptic membrane | 1.0E-02 | somatodendritic compartment | 6.7E-04 | cell leading edge | 7.7E-04 | sarcoplasm | 2.9E-03 |
| CC | voltage-gated potassium channel complex | 4.2E-03 | glutamatergic synapse | 1.1E-02 | neuron to neuron synapse | 1.7E-03 | actin-based cell projection | 1.2E-03 | cell leading edge | 3.0E-03 |
| CC | cell junction | 4.4E-03 | leading edge membrane | 1.2E-02 | asymmetric synapse | 2.4E-03 | postsynapse | 1.3E-03 | junctional sarcoplasmic reticulum membrane | 4.3E-03 |
| CC | potassium channel complex | 5.0E-03 | cell tip | 1.3E-02 | postsynaptic density | 2.4E-03 | cell cortex | 2.7E-03 | chromosomal region | 5.4E-03 |
| CC | BORC complex | 7.1E-03 | postsynaptic cytoskeleton | 1.4E-02 | neuron projection | 2.5E-03 | adherens junction | 3.1E-03 | cell junction | 6.5E-03 |
| CC | axolemma | 7.7E-03 | eukaryotic translation initiation factor 3 complex, eIF3m | 1.5E-02 | main axon | 6.3E-03 | lamellipodium | 3.2E-03 | postsynapse | 6.9E-03 |
| **Molecular Functions (MF)** |  |  |  |  |  |  |  |  |  |  |
| MF | phospholipase binding | 1.7E-03 | lysine-acetylated histone binding | 4.7E-04 | ubiquitin conjugating enzyme binding | 2.4E-03 | transcription factor binding | 2.4E-04 | protein serine kinase activity | 9.2E-04 |
| MF | efflux transmembrane transporter activity | 3.7E-03 | acetylation-dependent protein binding | 4.7E-04 | cytoskeletal anchor activity | 7.2E-03 | enzyme binding | 3.0E-04 | cell adhesion molecule binding | 1.9E-03 |
| MF | GTPase regulator activity | 7.6E-03 | transcription coregulator binding | 1.1E-03 | 1-phosphatidylinositol-4-phosphate 3-kinase activity | 8.5E-03 | DNA-binding transcription factor binding | 4.2E-04 | beta-catenin binding | 4.6E-03 |
| MF | nucleoside-triphosphatase regulator activity | 7.6E-03 | proline-rich region binding | 1.2E-03 | 1-phosphatidylinositol-3-kinase activity | 8.5E-03 | transcription coregulator activity | 7.6E-04 | phosphatidylinositol-3,4,5-trisphosphate binding | 5.8E-03 |
| MF | demethylase activity | 8.4E-03 | prostaglandin E receptor activity | 5.8E-03 | poly-pyrimidine tract binding | 1.0E-02 | RNA polymerase II-specific DNA-binding transcription factor binding | 8.5E-04 | transferase activity, transferring phosphorus-containing groups | 5.8E-03 |
| MF | bHLH transcription factor binding | 9.2E-03 | prostaglandin receptor activity | 6.1E-03 | ephrin receptor binding | 1.1E-02 | protein tyrosine kinase activity | 9.8E-04 | phosphatidylinositol-3,5-bisphosphate binding | 7.0E-03 |
| MF | histone demethylase activity | 1.1E-02 | transmembrane receptor protein tyrosine kinase activity | 8.3E-03 | branched-chain amino acid transmembrane transporter activity | 1.2E-02 | kinase activity | 1.0E-03 | cysteine-type endopeptidase regulator activity involved in apoptotic process | 7.7E-03 |
| MF | protein demethylase activity | 1.1E-02 | bicarbonate transmembrane transporter activity | 9.2E-03 | poly(U) RNA binding | 1.2E-02 | ATP binding | 1.9E-03 | histone H3K9 methyltransferase activity | 9.2E-03 |
| MF | RS domain binding | 1.1E-02 | transcription coactivator binding | 9.8E-03 | scaffold protein binding | 1.3E-02 | transcription coregulator binding | 1.9E-03 | ubiquitin-like protein transferase activity | 9.2E-03 |
| MF | transcription coregulator activity | 1.3E-02 | modification-dependent protein binding | 1.3E-02 | ubiquitin-like protein conjugating enzyme binding | 1.4E-02 | adenyl nucleotide binding | 2.7E-03 | cadherin binding | 9.5E-03 |

## Discussion

We performed the first systematic comparison of DNAm-health associations at two different time points during development (birth and childhood) on child outcomes spanning mental and physical domains, by jointly reanalyzing published multi-cohort EWAS meta-analyses from the PACE Consortium. Our findings lend three important new insights: 1. Effect sizes tend to be larger when DNAm is measured in childhood compared to at birth; 2. Even though EWAS effect sizes consistently increase over time for all outcomes examined, this did not necessarily lead to more significant findings; 3. DNAm signals are largely distinct between time points, but they correlate across outcomes, indicating shared associations with child health.

### Insight 1. EWAS effect sizes increase over time for all child health outcomes

Our first key finding is that across *all* five outcomes, mean EWAS effect sizes increased over time; i.e., they were stronger in the cross-sectional childhood analyses as compared to the prospective birth analyses. This may be due to a number of reasons: (i) the temporal proximity of the cross-sectional EWASs may better reflect immediate causal effects of DNAm on an outcome; (ii) in addition to genetic and prenatal environmental factors captured by DNAm at birth, DNAm in childhood may also reflect the accumulation of relevant postnatal environmental exposures and genetic effects^11^; (iii) peripheral blood (in childhood) may be a more informative tissue than cord blood (at birth), e.g. due to tissue differences in cell-type composition or immune profile – although we do not find evidence of enrichment for blood tissue-specific pathways in health-relevant CpGs that changed over time; and (iv) there may be unmeasured confounding (e.g., lifestyle, allergens) and reverse causation in childhood, which is more likely to affect cross-sectional analyses than prospective analyses.^19^ Indeed, Mendelian randomization studies suggest that for at least some sites, DNAm levels are a consequence, rather than a cause, of BMI^20,21^ or asthma^22^. While we can only speculate as to the most likely reason for the observed effect size increase, we can conclude that it is consistent for different outcomes, and to a comparable degree, hinting at potentially common driving factors.

### Insight 2. Higher effect sizes ≠ more significant findings

While EWAS effect sizes robustly increased, this did not necessarily result in more significant findings, as the signal also became ‘noisier’ with larger SE in childhood analyses. For BMI, the increase in effect sizes did correspond with an increase in statistical significance; however, for the other four outcomes significance on average either remained the same or actually decreased from birth to childhood, as evidenced most clearly for ADHD. Model specification is unlikely to explain differences in error, as outcome definitions and covariates were largely the same between the prospective and cross-sectional EWAS analyses; with the main differences relating to the predictor; i.e. when DNAm was assessed and cell-type proportion estimates (using different age- and tissue-appropriate reference panels).

Three other plausible ‘culprits’ for the noisier signal include sample size differences, between-study heterogeneity, and increasing DNAm variance with age. First, an imbalance in sample sizes (and associated power) between the birth and childhood EWASs could have led to differences in mean statistical significance. However, results remained largely consistent when re-running analyses restricting sample sizes to be equal between time points, ruling out this explanation. Second, we found that for all outcomes except GPF, between-study heterogeneity (i.e. systematic variability in effect sizes across the contributing cohorts) increased when DNAm was measured in childhood, potentially leading to more statistical uncertainty. Contributing factors may include (i) differences in DNAm assessment age, which varied substantially less in EWAS analyses at birth (cohort differences in the order of days) compared to EWAS in childhood (with age ranging from 5 to 17 years for asthma); and (ii) environmental differences between the included cohorts, which may cumulatively affect DNAm patterns (e.g., dietary factors, pollutant exposure, etc.), leading to more context-dependent associations in childhood. Importantly, however, between-study heterogeneity does not seem to fully account for increasing error in EWAS estimates over time. Indeed, when we re-ran meta-regression analyses only in ALSPAC, we found largely the same pattern of findings as the overall meta-analyses, meaning that sources of variability related to the use of multiple cohorts are unlikely to fully explain the observed temporal differences in EWAS signal.

A third explanation relates to DNAm variance. Variance for most DNAm sites increases with age (on average increasing 1.26 fold per year from birth), with only a minority of DNAm sites showing significant decreases in variance.^23^ It is likely that this increased variance reflects in part variation relevant to the studied health outcomes, e.g. reflecting additional important postnatal exposures, which results in increased effect sizes. At the same time, the increased variance likely also includes a substantial amount of variance unrelated to the studied health-related outcomes, increasing the standard error (i.e. adding noise) of the DNAm estimates and lowering power.

In summary, our findings caution against the assumption that larger effect sizes in EWAS lead to the identification of more hits. Rather, they suggest that statistical power varies depending on factors such as the degree of uncertainty and study heterogeneity, the timing of DNAm assessment, and the potentially causal nature and direction of associations between DNAm and a given outcome.

### Insight 3. Epigenetic signals associated with child outcomes are time-specific and pleiotropic

Our analyses correlating EWAS estimates between time points reveal largely distinct association signals at birth versus in childhood: generally, for a given outcome, estimates at birth did not correlate with those in childhood – or only modestly in the case of ADHD. Based on the available data, it is not possible to establish whether this specificity in DNAm signals extends more broadly to other life stages, or whether DNAm associations become more stable and comparable after some developmental point.^10,23^ These temporal differences raise the question of which DNAm assessment time point may be most relevant for health. For biomarker purposes, our results suggest that DNAm estimates from cross-sectional childhood analyses may explain higher phenotypic variance, but at the cost of higher uncertainty of estimates. This may lead to noisier methylation profile scores (MPS; akin to polygenic scores or PGS), which are also more likely to reflect consequences of a phenotype, and thus less useful for prediction of later outcomes.^24^ Our results caution that MPS developed from one DNAm time point may generalize poorly to different time-points. As such, repeated assessments of DNAm and the combination of multiple age-specific scores may be needed to improve MPS performance, although specific guidelines are difficult to formulate based on the present findings. For instance, MRSs based on allergy-related EWAS performed similarly well when tested at both age 6 and 10 years,^25^ but differences between birth and childhood methylation profiles are likely more impactful.

Surprisingly, the consistency of estimates across some child outcomes was larger than between time points for the same outcome. Our correlation analyses suggest that DNAm associations with ADHD, GPF and asthma are to some degree shared. This is in line with previous studies pointing to phenotypic and genetic correlations between these outcomes^26–29^, and may point towards an early shared origin of these conditions reflected in the methylome or network effects among the phenotypes. Enrichment analyses suggest that neural pathways may be involved in all tested health outcomes (particularly mental phenotypes) and may partly explain the observed correlation. On the other hand, the observed negative correlation between (birth/childhood) ADHD and (childhood) BMI estimates is more perplexing. Children with ADHD are more likely to be overweight and vice versa,^30,31^ and BMI and ADHD also show positive genetic correlations.^27,32^ This may indicate that epigenetic risk mechanisms for ADHD are associated with lower BMI in childhood, but are overshadowed by (non-methylation related) mechanisms causing positive phenotypic correlations.

### Study limitations

Our meta-regression approach enabled us to quantify longitudinal trends (changes in EWAS effect sizes, standard error and statistical significance) in the relationship between DNAm assessed at multiple time points (birth, childhood) and various child health outcomes, as well as to estimate how EWAS signals correlate over time and across outcomes. However, summary statistics-based approaches also have several limitations. While we accounted for repeated measures from the same cohorts, the degree of sample overlap across time points and outcomes could not be explicitly modeled based on summary data.^33^ That said, sensitivity analyses in a single cohort with largely overlapping samples did not alter conclusions. In addition, modeling effect size changes and between-study heterogeneity more granularly to incorporate information on specific age of DNAm assessment (i.e. [gestational] age at birth or childhood) would also require individual-level analyses. Another individual characteristic we cannot model with the given data is sex. It is possible that associations may differ depending on sex and affect childhood DNAm associations disproportionately, especially after puberty. Future studies with individual-level data should also study the impact of increasing DNAm variance on association estimates. Lastly, we could not perform formal epigenetic correlation tests on individual-level data, and the correlation analyses of regression coefficients should therefore be interpreted as hypothesis-generating for future research.

With current study designs, it is also impossible to disentangle timing differences from tissue differences between cord and peripheral blood. Each EWAS adjusted for estimated cell proportions, but such corrections only adjust for cell composition differences within time points and tissues, but not across tissues^26^. Morphogenesis pathways showed some evidence of enrichment in case of BMI and asthma, leaving the possibility open for an involvement of tissue differentiation. Future studies are needed that examine different tissues at birth (to determine the specificity of the findings to cord blood as opposed to the neonatal period in general) as well as DNAm at multiple time points in childhood (to test if effect sizes change non-linearly across developmental periods).^10^ While our analyses provided important new insights into genome-wide trends, they were mostly underpowered to identify *specific* DNAm sites showing longitudinal changes in associations at a genome-wide level of significance; as such, larger studies are needed to reliably characterize epigenetic changes in associations for individual sites. Finally, expanding analyses to other outcomes should be pursued in future research.

### Conclusions

Overall, our results suggest developmentally-specific associations between DNAm and child health outcomes, when assessing DNAm at birth vs childhood. This implies that EWAS results from one time point are unlikely to generalize to another (at least based on birth vs childhood comparisons): a consequential finding, given that most research to date examines DNAm at a single assessment time-point. Longitudinal studies with repeated epigenetic assessments are direly needed to shed light on the dynamic relationship between DNAm, development and health, as well as to enable the creation of more reliable and generalizable epigenetic biomarkers. More broadly, this study underscores the importance of considering the time-varying nature of DNAm in epigenetic research and supports the potential existence of epigenetic ‘timing effects’ on child health.

## Methods

### Data

We requested cohort-level epigenome-wide summary statistics from five meta-analytic studies previously performed by the PACE Consortium. We obtained permission for re-analysis from the meta-analysis leads and representatives of all originally participating cohorts, except for the GOYA study, which was excluded here from further analysis. The Erasmus MC Medical Ethics Review Committee and respective local ethics committees previously approved the included studies.^7–9,17,18^

EWAS summary statistics included the association between DNAm (predictor) and a phenotype (outcome). DNAm was either measured in cord blood at birth or in peripheral blood in childhood with either Illumina 450K or EPIC arrays (although only 450K DNAm sites remained after QC, see below). Predictors were the DNAm betas ranging from 0 to 1, corresponding to 0 to 100 percent methylation, with analyses for GPF and Sleep having trimmed DNAm outliers outside the range of [25^th^ percentile - (3*interquartile range (IQR) to 75^th^ percentile + 3*IQR). ADHD, GPF and sleep were assessed via parental questionnaires and BMI was computed based on measured height and weight. ADHD, GPF and sleep were assessed via parental questionnaires and BMI was computed based on measured height and weight. These were modelled as continuous measures that were z-score standardized within each cohort.^8,9,17,18^ Asthma was classified based on doctor’s diagnosis and symptoms in past years, and analyzed in a dichotomous fashion.^7^ All EWAS were adjusted for sex, maternal age, maternal education, maternal smoking, cell proportions and possible batch effects, in addition to other variables, which differed depending on outcome and time-point. A variety of analysis models were employed, such as OLS linear models (ADHD, sleep), robust linear models (GPF, BMI) and logistic regression (asthma). For ADHD, some cohorts applied linear mixed models, when repeated measures of ADHD were available (Table 1).

We applied the following additional quality control: 1. Kept only autosomal DNAm sites, 2. removed DNAm sites with information in less than three cohorts or 1000 participants per time-point, 3. kept only CpG sites present both at birth and in childhood, 4. removed cross-reactive probes using the maxprobe 0.0.2 package (https://github.com/markgene/maxprobes). Finally, to examine whether the differences in statistical significance were influenced by sample size differences, we also performed sensitivity analyses with similar sample sizes at both time points. We removed (combination of) cohorts which resulted in the most similar sample sizes between cohorts (Table S1).

### Statistical Analysis

Each summary statistic contained information on the regression coefficient (β_jk_) and standard error. β represents the expected difference in the outcome in SD between no and full methylation at the tested CpG site at DNAm assessment time-point j (birth or childhood) estimated in cohort k. We applied multi-level meta-regressions to pool effect sizes across cohorts and to model changes in effect sizes depending on DNAm assessment time-point. Repeated measures from cohorts that contributed association estimates for both DNAm at birth and in childhood were taken into account with a random intercept. The main model therefore took the form of:

β_jk=_β_birth_+β_Δchildhood_+u_k_+r_k_

β_birth_ is the intercept and represents the pooled variance-weighted associations of methylation at a CpG site on an outcome at birth or childhood, respectively.

β_Δchildhood_ refers to the change in association from DNAm at birth to childhood.

u_k_ is the study random effect and refers to deviation of the mean associations within cohort k from overall mean associations.

r_k_ denotes residual error

We also ran a statistically identical model with reverse time direction to extract DNAm effects at childhood. We applied these meta-regression models to each DNAm site separately using metafor 4.2.0^34^ in R 4.2.2^35^. After estimating the associations and their change for each CpG site, we aggregated statistics across the genome to characterize global trends. Specifically, we examined across all CpG sites the mean absolute effect size at birth (|β̅_birth_|), mean absolute effect size in childhood (|β̅_childhood_|), and the mean effect size difference between birth and childhood (|Δβ̅_Δchildhood_|). In addition, we examined trends of statistical significance by taking the mean z test statistic of β_birth_ (|z̄_birth_|) and β_childhood_ (|z̄_childhood_|), representing the evidence of association for DNAm at birth and childhood, respectively. Furthermore, we also characterized the change in mean statistical significance from birth to childhood methylation (|Δz̄|).

We also examined whether between-study heterogeneity changed between birth and childhood estimates by adding a random slope of β_Δchildhood_ on the cohort level. We extracted τ, which indicates to which degree DNAm effects vary due to between-study heterogeneity within 1SD. In other words, assuming no sampling error and normal distribution, 67% of estimates are expected to be within β+-τ due to study differences. Reported correlations are spearman correlations. GO term enrichment for DNAm sites with nominally significant change and nominally significant association for at least one time point was tested using missMethyl 1.36.0.^36,37^

## Data Availability

Original analysis code and example data can be found at https://github.com/aneumann-science/epigenetic_timing_effects. Full meta-analysis summary statistics can be downloaded at https://doi.org/10.5281/zenodo.10720466 (Will be made public after acceptance). For individual cohort summary statistics or individual-level data, please see original publications for details on how to request them.
1. Neumann, A. et al. Association between DNA methylation and ADHD symptoms from birth to school age: a prospective meta-analysis. Transl. Psychiatry 10, 398 (2020).
2. Rijlaarsdam, J. et al. DNA methylation and general psychopathology in childhood: an epigenome-wide meta-analysis from the PACE consortium. Mol. Psychiatry 28, (2023).
3. Sammallahti, S. et al. Longitudinal associations of DNA methylation and sleep in children: a meta-analysis. Clin. Epigenetics 14, 83 (2022).
4. Vehmeijer, F. O. L. et al. DNA methylation and body mass index from birth to adolescence: meta-analyses of epigenome-wide association studies. Genome Med. 12, 105 (2020).
5. Reese, S. E. et al. Epigenome-wide meta-analysis of DNA methylation and childhood asthma. J. Allergy Clin. Immunol. 143 (2019).

https://github.com/aneumann-science/

## Acknowledgments

We thank all the children and families who took part in the original study cohorts, as well as the support of hospitals, midwives, and pharmacies. We also gratefully acknowledge all researchers who contributed to the original PACE meta-analyses included in this study.

The work of AN and CAMC was supported by the European Research Council (TEMPO; grant agreement No 101039672) and the European Union’s HorizonEurope Research and Innovation Programme (FAMILY, grant agreement No 101057529). The work of CAMC is further supported by the European Union’s Horizon 2020 Research and Innovation Programme (EarlyCause, grant agreement No 848158; HappyMums, grant agreement No 101057390). CAMC and JFF are supported by the European Union’s Horizon Europe Programme (STAGE project, grant agreement no.101137146). This research was conducted while CAMC was a Hevolution/AFAR New Investigator Awardee in Aging Biology and Geroscience Research.

LST was supported by the CAPICE (Childhood and Adolescence Psychopathology: unravelling the complex etiology by a large Interdisciplinary Collaboration in Europe) project, the European Union’s Horizon 2020 research and innovation programme, Marie Sklodowska Curie Actions – MSCA-ITN-2016 – Innovative Training Networks under grant agreement number 721567.

SJL was supported in part by the Intramural Research Program of the NIH, NIEHS Z01 ES49019.

SEH and CMP was partly supported by The Norwegian Research council no 262700 and the Norwegian Cancer Society project no 244291.

See supplementary information for cohort-specific acknowledgments and funding.

Support for title page creation and format was provided by AuthorArranger, a tool developed at the National Cancer Institute.

## Author contributions

AN and CC developed the study design and drafted the manuscript. AN performed statistical analysis. CC supervised the study. All co-authors contributed to the original epigenome-wide association studies used in the meta-regression and revised the manuscript critically.

