## Supplementary figures and images for "Epigenetic timing effects on child developmental outcomes: A longitudinal meta-regression of findings from the Pregnancy And Childhood Epigenetics Consortium"

### Figure S1

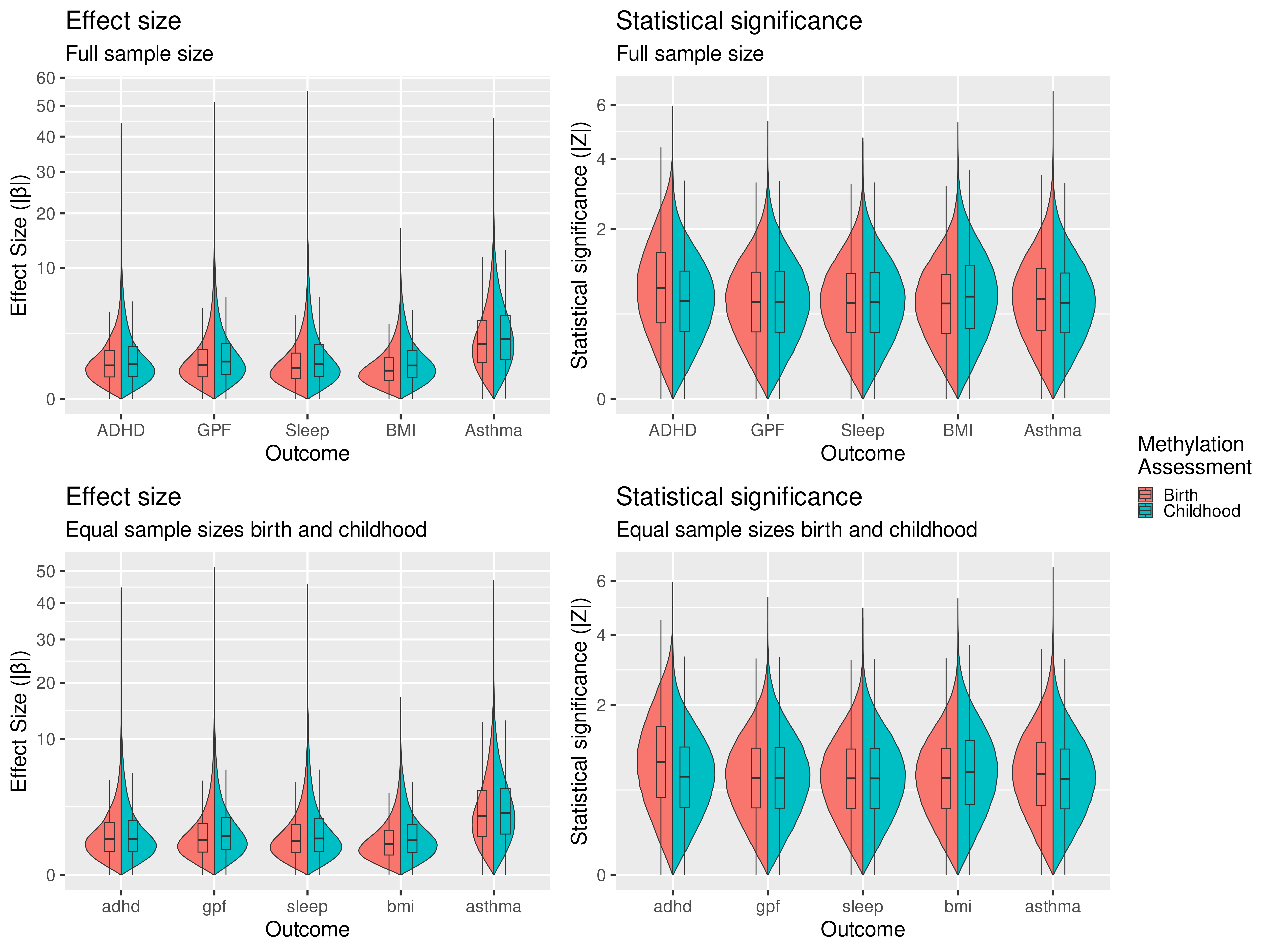

### Figure S2

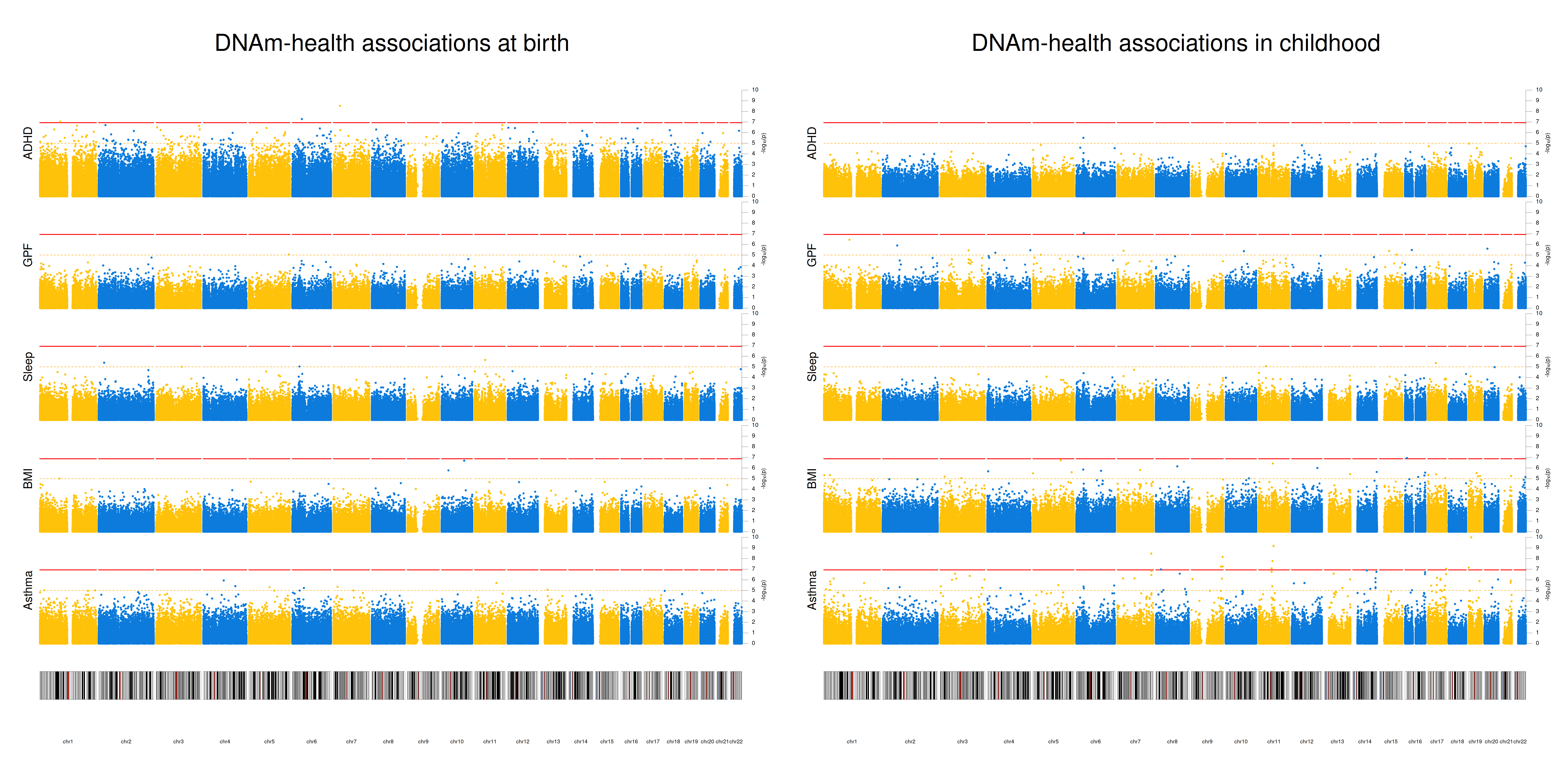

### Figure S3

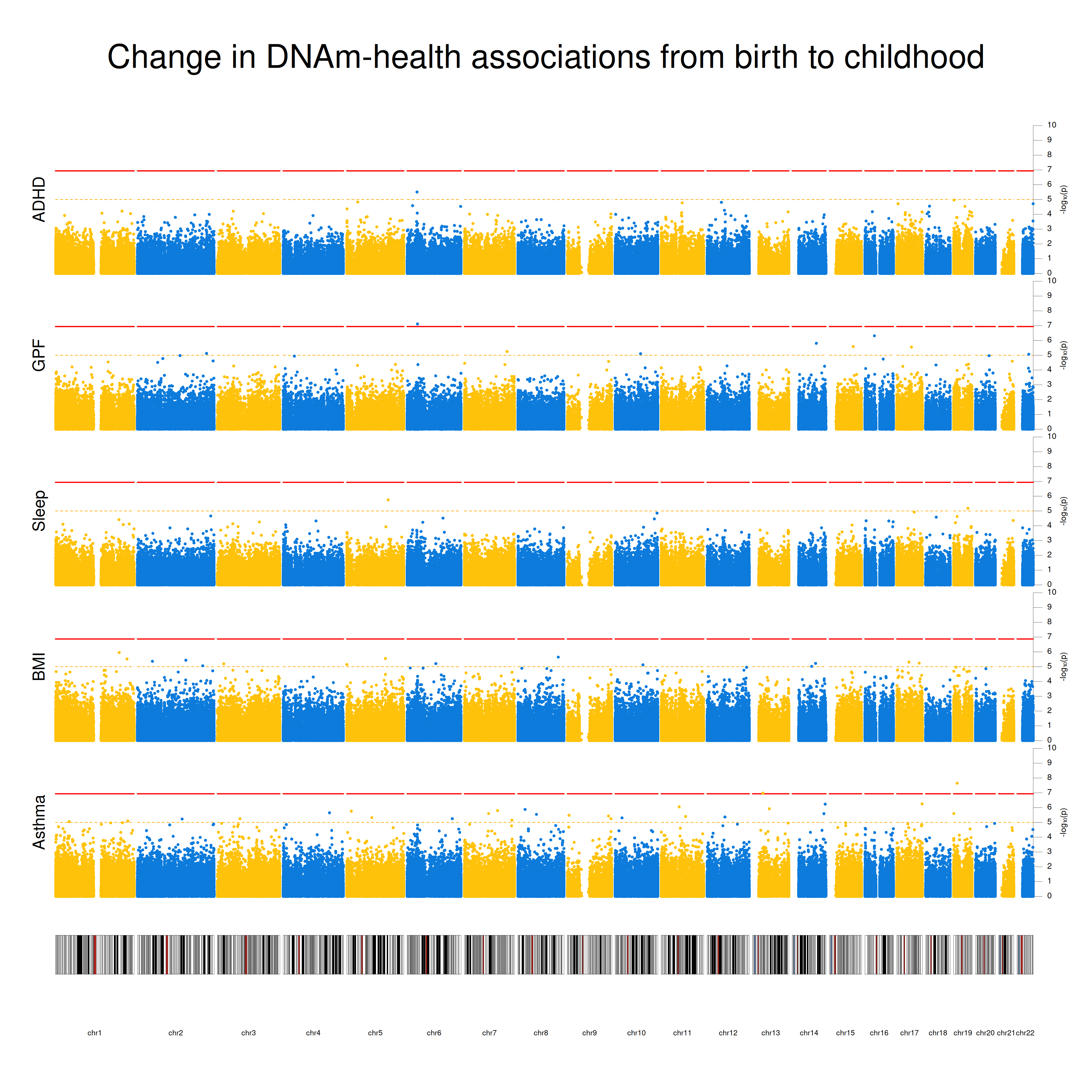

### Figure S4

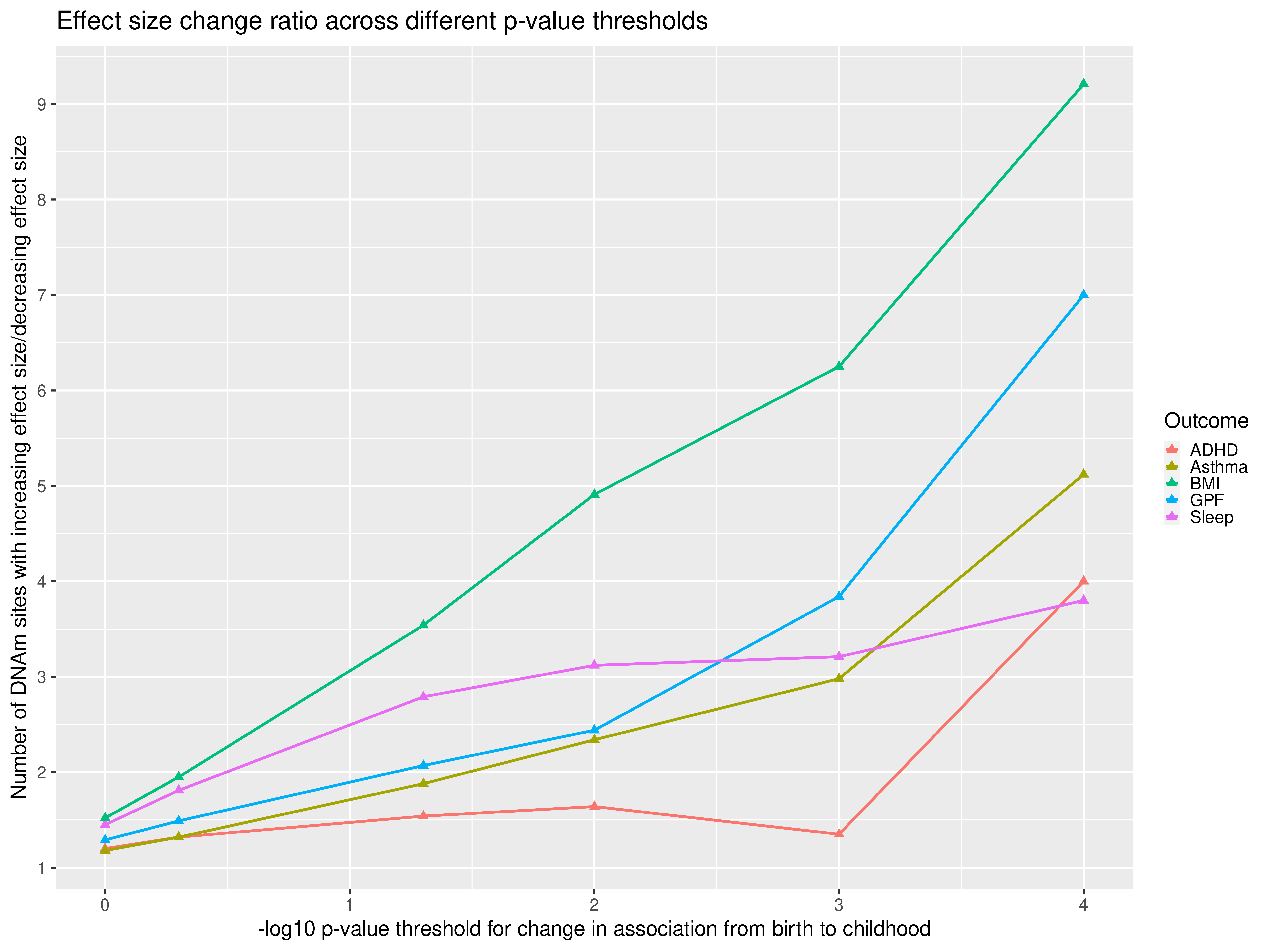

### Figure S5

#### β correlations between time points and outcomes (ALSPAC only)

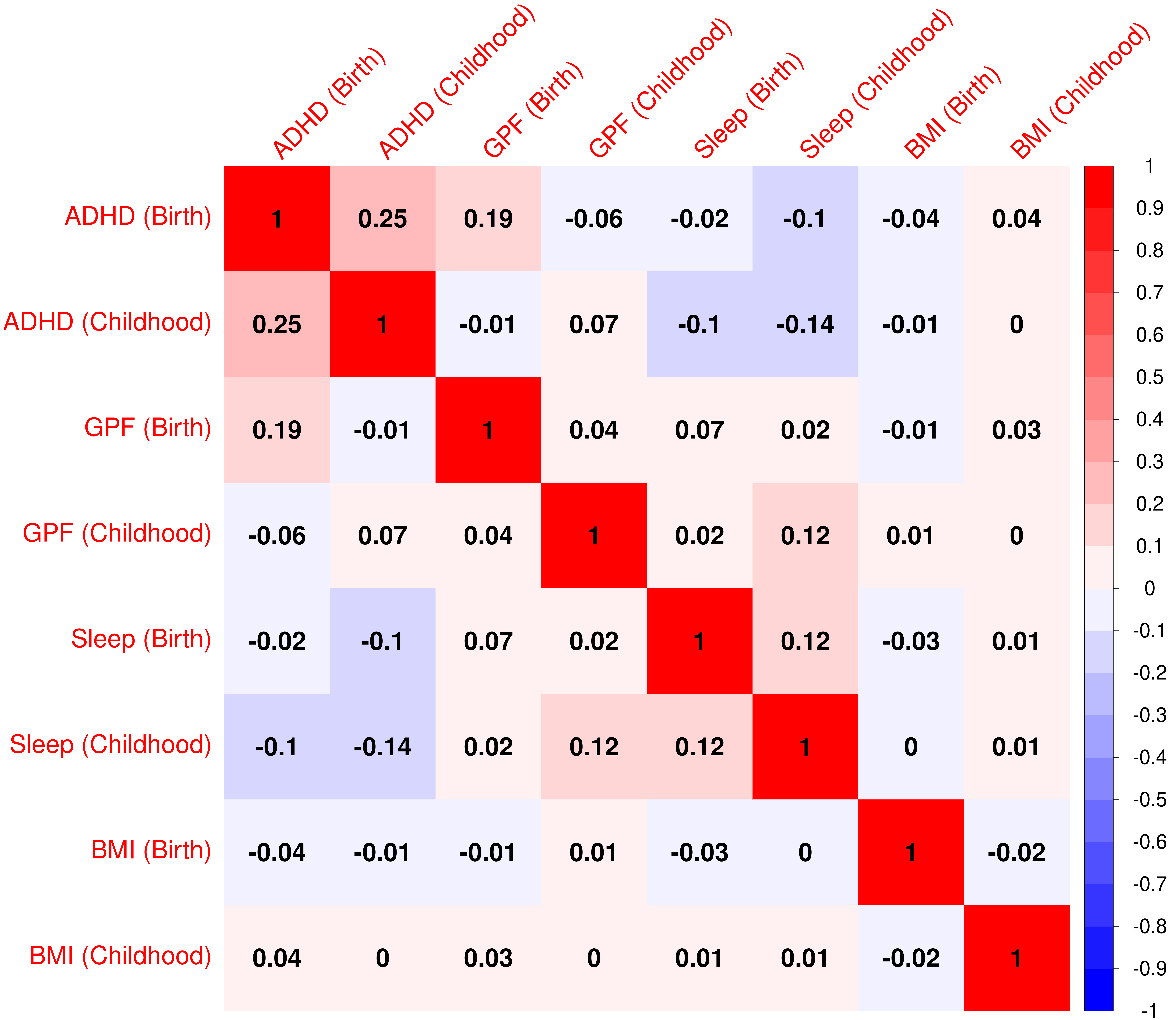
