## Supplementary Information for "Epigenetic timing effects on child developmental outcomes: A longitudinal meta-regression of findings from the Pregnancy And Childhood Epigenetics Consortium"

### Supplement:
### Cohort-specific information

Following funding statements are adapted from the original PACE meta-analyses included in this study.[1–5]

**Avon Longitudinal Study of Parents and Children (ALSPAC):**

Pregnant women resident in Avon, UK with expected dates of delivery between 1st April 1991 and 31st December 1992 were invited to take part in the study. 20,248 pregnancies have been identified as being eligible and the initial number of pregnancies enrolled was 14,541. Of the initial pregnancies, there was a total of 14,676 foetuses, resulting in 14,062 live births and 13,988 children who were alive at 1 year of age. The total sample size for analyses using any data collected after the age of seven is therefore 15,447 pregnancies, resulting in 15,658 foetuses. Of these 14,901 children were alive at 1 year of age.

See following references for full cohort descriptions:

Boyd A, Golding J, Macleod J, Lawlor DA, Fraser A, Henderson J, Molloy L, Ness A, Ring S, Davey Smith G. Cohort Profile: The 'Children of the 90s'; the index offspring of The Avon Longitudinal Study of Parents and Children (ALSPAC). International Journal of Epidemiology 2013; 42: 111-127.

Fraser A, Macdonald-Wallis C, Tilling K, Boyd A, Golding J, Davey Smith G, Henderson J, Macleod J, Molloy L, Ness A, Ring S, Nelson SM, Lawlor DA. Cohort Profile: The Avon Longitudinal Study of Parents and Children: ALSPAC mothers cohort. International Journal of Epidemiology 2013; 42:97-110.

Please note that the study website contains details of all the data that is available through a fully searchable data dictionary and variable search tool" and reference the following webpage: http://www.bristol.ac.uk/alspac/researchers/our-data

Ethical approval for the study was obtained from the ALSPAC Ethics and Law Committee and the Local Research Ethics Committees. Consent for biological samples has been collected in accordance with the Human Tissue Act (2004). Informed consent for the use of data collected via questionnaires and clinics was obtained fromparticipants following the recommendations of the ALSPAC Ethics and Law Committee at the time.

We are extremely grateful to all the families who took part in this study, the midwives for their help in recruiting them, and the whole ALSPAC team, which includes interviewers, computer and laboratory technicians, clerical workers, research scientists, volunteers, managers, receptionists and nurses.

The UK Medical Research Council and Wellcome (grant ref: 217065/Z/19/Z) and the University of Bristol provide core support for ALSPAC. Methylation data in the ALSPAC cohort were generated as part of the UK BBSRC funded (grant numbers: BB/I025751/1 and BB/I025263/1) Accessible Resource for Integrated Epigenomic Studies (ARIES, http://www.ariesepigenomics.org.uk). D.C., work in a Unit that is supported by the University of Bristol and the UK Medical Research Council (grant number: MC_UU_00011/5). This publication is the work of the authors and Alexander Neumann will serve as guarantors for the contents of this paper

**BAMSE**

BAMSE was supported by The Swedish Research Council, The Swedish Heart-Lung Foundation and Region Stockholm (ALF and database maintenance)

**CHAMACOS**

CHAMACOS cohort research was supported by grants from the National Institute of Environmental Health Science (NIEHS) [P01 ESO09605, 5UG30D023356, R01ES012503, R01ES021369, R01ES023067, R24ES0285529]; Environmental Protection Agency (RD83273401, RD83171001), and the JPB Foundation. Its contents are solely the responsibility of the authors and do not necessarily represent the official views of NIEHS, EPA, or JPB Foundation.

We are grateful to the laboratory and field staff and participants of the CHAMACOS study for their contributions. We are thankful to Hong Quach who helped with 450K methylation analyses.

**CHOP**

The CHOP study has been carried out with partial financial support from the Commission of the European Community, specific RTD Programme "Quality of Life and Management of Living Resources", within the Fifth Framework Program (research grants no. QLRT-2001-00389 and QLK1-CT-200230582), the Sixth Framework Program (contract no. 007036), and Seventh Framework Programme (EarlyNutrition; grant agreement no. 289346), the EU H2020 project LIFECYCLE under grant no. 733206 and the European Research Council Advanced Grant META-GROWTH (ERC-2012-AdG – no.322605) and with financial support from Polish Ministry of Science and Higher Education (2571/7.PR/2012/2).

We like to thank the participating families and all project partners for their enthusiastic support of the project work. We also like to thank Dr Eva Reischl and team at the Genome Analysis Center of Helmholtz Zentrum Muenchen, Germany for DNA extraction, bisulfite conversion and methylation analysis. We also like to acknowledge The European Childhood Obesity Trial Study Group for their continuous and salient support of the CHOP project: Philippe Goyens, Clotilde Carlier, Joana Hoyos, Pascale Poncelet, and Elena Dain (Universite Libre de Bruxelles – (ULB) –Brussels , Belgium); Jean-Noel Van Hees (CHC St Vincent– Françoise Martin, Annick Xhonneux, Jean-Paul Langhendries, and Jean-Noel Van Hees - Liège-Rocourt, Belgium); Ricardo Closa-Monasterolo, Joaquin Escribano, Veronica Luque, Georgina Mendez, Natalia Ferre, and Marta Zaragoza-Jordana (Universitat Rovira i Virgili, Institut d'Investigacio´ Sanitaria Pere Virgili, Taragona, Spain); Marcello Giovannini, Enrica Riva, Carlo Agostoni, Silvia Scaglioni, Elvira Verduci, Fiammetta Vecchi, and Alice Re Dionigi (University of Milano, Milano, Italy); Jerzy Socha, Piotr Socha and Anna Stolarczyk (Children's Memorial Health Institute, Department of Gastroenterology, Hepatology and Immunology, Warsaw, Poland); Anna Dobrzanska and Dariusz Gruszfeld (Children's Memorial Health Institute, Neonatal Intensive Care Unit, Warsaw, Poland); Roman Janas (Children's Memorial Health Institute, Diagnostic Laboratory, Warsaw, Poland); Emmanuel Perrin (Danone Research Centre for Specialized Nutrition, Schiphol, the Netherlands); Rudiger von Kries (Division of Pediatric Epidemiology, Institute of Social Pediatrics and Adolescent Medicine, Ludwig Maximilians University of Munich, Munich, Germany); Helfried Groebe, Anna Reith, and Renate Hofmann (Klinikum Nurnberg Sued, Nurnberg, Germany); and Berthold Koletzko, Veit Grote, Martina Weber, Peter Rzehak, Sonia Schiess, Jeannette Beyer, Michaela Fritsch, Uschi Handel, Ingrid Pawellek, Sabine Verwied-Jorky, Iris Hannibal, Hans Demmelmair, Gudrun Haile, and Melissa Theurich (Division of Nutritional Medicine and Metabolism, Dr von Hauner Childrens Hospital, Ludwig-Maximilians Universität München (LMU), Munich, Germany).

**CHR**

The Saguenay-Lac-Saint-Jean asthma familial cohort was supported by Laprise grants from the Canadian Institute of Health Research (CIHR).

**CHS**

This work was supported by NIEHS grants K01ES017801, R01ES022216, and P30ES007048.

We are indebted to the school principals, teachers, students and parents in each of the study communities for their cooperation and especially to the members of the health testing field team for their efforts. We would like to express our sincere gratitude to Steve Graham and Robin Cooley at the California Biobank Program and Genetic Disease Screening Program within the California Department of Public Health for their assistance and advice regarding newborn bloodspots. The biospecimens and/or data used in this study were obtained from the California Biobank Program, (SIS request number(s) 479)" Section 6555(b), 17 CCR. The California Department of Public Health is not responsible for the results or conclusions drawn by the authors of this publication.

**DOMINO**

The DOMInO Study and 3 and 5 year follow-up of the DOMInO children were supported by grants (349301, 570109) and Fellowships (APP1004211, APP1046207, APP1061074 and APP1052388) from the National Health and Medical Research Council of Australia (NHMRC). The epigenetic analyses of DOMInO samples were supported by the Science and Industry Endowment Fund (RP03-064), Diabetes Australia Research Trust and National Institutes of Health grant R35 CA 209859.

The authors acknowledge the DOMInO Steering committee for their support of this study (M Makrides, RA Gibson, AJ McPhee, LN Yelland, J Quinlivan, K Best, P Ryan). We would like to thank the families who participated; the medical, nursing, and research staff in each participating center and the whole DOMInO team, including the staff of the Child Nutrition Research Centre; and the staff of the Data Management and Analysis Centre, University of Adelaide, Adelaide, Australia. We gratefully acknowledge Susan van Dijk for her role in co-ordinating the DNA methylation analyses on the participants.

**DCSH**

The Drakenstein Child Health Study (DCHS) was funded by the Bill & Melinda Gates Foundation (OPP 1017641, OPP1017579), Medical Research Council South Africa, and the National Research Foundation South Africa. Additional support for the DNA methylation work was by the Eunice Kennedy Shriver National Institute of Child Health and Human Development of the National Institutes of Health (NICHD) under Award Number R21HD085849, and the Fogarty International Center (FIC).

The authors thank the study and clinical staff at Paarl Hospital, Mbekweni and TC Newman clinics, as well as the CEO of Paarl Hospital, and the Western Cape Health Department for their support of the study. The authors thank the families and children who participated in this study. The authors also thank Dr. Michael S. Kobor and his team at the University of British Columbia for the generation, pre-processing and quality control of the DNA methylation data (data generation: Julia L MacIsaac, David TS Lin, Katia E Ramadori; pre-processing/quality control: Nicole Gladish).

**EDEN**

EDEN: Foundation for Medical Research (FRM), National Agency for Research (ANR), National Institute for Research in Public Health (IRESP: TGIR cohorte santé 2008 program), French Ministry of Health (DGS), French Ministry of Research, Inserm Bone and Joint Diseases National Research (PRO-A) and Human Nutrition National Research Programs, Paris–Sud University, Nestlé, French National Institute for Population Health Surveillance (InVS), French National Institute for Health Education (INPES), the European Union FP7 programmes (FP7/2007-2013, HELIX, ESCAPE, ENRIECO, MEDall projects), Diabetes National Research Program (through a collaboration with the French Association of Diabetic Patients (AFD)), French Agency for Environmental Health Safety (now ANSES), MutuelleGénérale de l'EducationNationale (MGEN), French National Agency for Food Security, Health and Environment-wide Associations based on Large population Surveys (HEALS) and the French-speaking association for the study of diabetes and metabolism (ALFEDIAM).

We are indebted to all the children and their parents for participation, as well as to the research nurses, research assistants, and laboratory personnel involved in the EDEN study. WE are also greatful to the the EDEN Mother–Child Cohort Study Group which includes: I. Annesi-Maesano, J.Y Bernard, J. Botton, M.A. Charles, P. Dargent-Molina, B. de Lauzon-Guillain, P. Ducimetière, M. de Agostini, B. Foliguet, A. Forhan, X. Fritel, A. Germa, V. Goua, R. Hankard, B. Heude, M. Kaminski, B. Larroque†, N. Lelong, J. Lepeule, G. Magnin, L. Marchand, C. Nabet, F. Pierre, R. Slama, M.J. Saurel-Cubizolles, M. Schweitzer, O. Thiebaugeorges.

**GALA II**

GALA II: This work was supported in part by the Sandler Family Foundation, the American Asthma Foundation, the RWJF Amos Medical Faculty Development Program, Harry Wm. and Diana V. Hind Distinguished Professor in Pharmaceutical Sciences II, National Institutes of Health R01HL117004, R01HL128439, R01HL135156, X01HL134589, National Institute of Health and Environmental Health Sciences, R21ES24844, the National Institute on Minority Health and Health Disparities P60MD006902, U54MD009523, R01MD010443, and Tobacco-Related Disease Research Program under Award Number 24RT-0025.

**GECKO**

The GECKO Drenthe birth cohort was funded by an unrestricted grant of Hutchison Whampoa Ld, Hong Kong and supported by the University of Groningen, Well Baby Clinic Foundation Icare, Noordlease and Youth Health Care Drenthe. This methylation project in the GECKO Drenthe cohort was supported by the Biobanking and Biomolecular Research Infrastructure Netherlands (CP2011-19). This project received funding from the European Union's Horizon 2020 research and innovation programme (733206, LIFECYCLE).

We are grateful to the families who took part in the GECKO Drenthe birth cohort, the midwives, gyneacologists, nurses and GPs for their help for recruitment and measurement of participants, and the whole team from the GECKO Drenthe study.

Generation R Study (Generation R): The general design of the Generation R Study is made possible by financial support from the Erasmus Medical Center, Rotterdam, the Erasmus University Rotterdam, the Netherlands Organization for Health Research and Development and the Ministry of Health, Welfare and Sport. The EWAS data was funded by a grant from the Netherlands Genomics Initiative (NGI)/Netherlands Organisation for Scientific Research (NWO) Netherlands Consortium for Healthy Aging (NCHA; project nr. 050-060-810), by funds from the Genetic Laboratory of the Department of Internal Medicine, Erasmus MC, and by a grant from the National Institute of Child and Human Development (R01HD068437).

The Generation R Study is conducted by the Erasmus Medical Center in close collaboration with the School of Law and Faculty of Social Sciences of the Erasmus University Rotterdam, the Municipal Health Service Rotterdam area, Rotterdam, the Rotterdam Homecare Foundation, Rotterdam and the Stichting Trombosedienst & Artsenlaboratorium Rijnmond (STAR-MDC), Rotterdam. We gratefully acknowledge the contribution of children and parents, general practitioners, hospitals, midwives and pharmacies in Rotterdam. The study protocol was approved by the Medical Ethical Committee of the Erasmus Medical Centre, Rotterdam. Written informed consent was obtained for all participants. The generation and management of the Illumina 450K methylation array data (EWAS data) for the Generation R Study was executed by the Human Genotyping Facility of the Genetic Laboratory of the Department of Internal Medicine, Erasmus MC, the Netherlands. We thank Mr. Michael Verbiest, Ms. Mila Jhamai, Ms. Sarah Higgins, Mr. Marijn Verkerk and Dr. Lisette Stolk for their help in creating the EWAS database. We thank Dr. A.Teumer for his work on the quality control and normalization scripts.

**GLAKU**

Glycyrrhizin in Licorice (GLAKU): The study has been supported by Academy of Finland, University of Helsinki, Hope and Optimism Initiative, Finnish Foundation for Pediatric Research, Sigrid Juselius Foundation, Jalmari and Rauha Ahokas Foundation, Signe and Ane Gyllenberg Foundation, Yrjo Jahnsson Foundation, Juho Vainio Foundation, Emil Aaltonen Foundation, and Ministry of Education and Culture, Finland. The 352 samples were genotyped at the Genotyping and Sequencing Core Facility of the Estonian Genome Centre, University of Tartu.

We thank all the Glaku children and their parents for their enthusiastic participation. We also thank all the research nurses, research assistants, and laboratory personnel involved in the Glaku study.

**Healthy Start**

The Healthy Start study was supported by grants from the National Institute of Diabetes and Digestive and Kidney Diseases (R01DK076648), the National Institute of Environmental Health Sciences (R01ES022934), and the Office of the Director (UH3OD023248) of the National Institutes of Health. APS was additionally supported by a grant from the National Institute of Environmental Health Sciences (R00ES025817).

We would like to acknowledge the contributions of the study coordinator, Ms. Mercedes Martinez, MPH, the study staff, and the participating families for their continued involvement in the Healthy Start study.

**HELIX**

Human Early Life Exposome (HELIX) The research leading to these results has received funding from the European Community's Seventh Framework Programme (FP7/2007-206) under grant agreement no 308333—the HELIX project. INMA data collections were supported by grants from the Instituto de Salud Carlos III, CIBERESP, the Conselleria de Sanitat, Generalitat Valenciana, Department of Health of the Basque Government; the Provincial Government of Gipuzkoa, and the Generalitat de Catalunya-CIRIT. KANC was funded by the grant of the Lithuanian Agency for Science Innovation and Technology (6-04-2014_31V-66). The Norwegian Mother, Father and Child Cohort Study is supported by the Norwegian Ministry of Health and Care Services and the Ministry of Education and Research. The Rhea project was financially supported by European projects, and the Greek Ministry of Health (Program of Prevention of Obesity and Neurodevelopmental Disorders in Preschool Children, in Heraklion district, Crete, Greece: 2011–2014; 'Rhea Plus': Primary Prevention Program of Environmental Risk Factors for Reproductive Health, and Child Health: 2012–2015). The work was also supported by MICINN (MTM2015-68140-R) and Centro Nacional de Genotipado-CEGEN-PRB2-ISCIII. This project has received funding from the European Union's Horizon 2020 research and innovation programme under grant agreement No 874583.

We would like to thank all the children and their families for their generous contribution.

INMA

INfancia y Medio Ambiente (INMA) Main funding of the epigenetic studies, and of birth and nine years of age assessments/visits in INMA were grants from Instituto de Salud Carlos III (Red INMA G03/176, CB06/02/0041, CP18/00018, PI041436, PI081151 incl. FEDER funds, PI12/01890 incl. FEDER funds, CP13/00054 incl. FEDER funds), CIBERESP, Spanish Ministry of Health (FIS-PI04/1436, FIS-PI08/1151 including FEDER funds, FIS-PI11/00610, FIS-FEDER-PI06/0867, FIS-FEDER-PI03-1615), Spanish Ministry of Economy and Competitiveness (SAF2012-32991 incl. FEDER funds), Agence Nationale de Securite Sanitaire de l'Alimentation de l'Environnement et du Travail (1262C0010), Generalitat de Catalunya-CIRIT 1999SGR 00241, Generalitat de Catalunya-AGAUR (2009 SGR 501, 2014 SGR 822), Fundació La marató de TV3 (090430), EU Commission (261357-MeDALL: Mechanisms of the Development of ALLergy, 308333, 603794, and 634453), and European Research Council (268479-BREATHE: BRain dEvelopment and Air polluTion ultrafine particles in scHool childrEn). We acknowledge support from the Spanish Ministry of Science and Innovation and the State Research Agency through the "Centro de Excelencia Severo Ochoa 2019-2023" Program (CEX2018-000806-S), and support from the Generalitat de Catalunya through the CERCA Program.

The authors would particularly like to thank all the participants for their generous collaboration. A full roster of the INMA Project Investigators can be found at http://www.proyectoinma.org/presentacion-inma/listado-investigadores/en_listado-investigadores.html.

**IOWF1 and F2**

The IoW 1989 (IOW F1) cohort was supported by the National Institute of Allergy and Infectious Diseases under award numbers R01 AI091905 (PI: Wilfried Karmaus), R01 AI061471 (PI: Susan Ewart), and R01 AI121226 (PIs: Zhang, Holloway). The 18-year follow-up by a grant from the National Heart and Blood Institute (R01 HL082925, PI: S. Hasan Arshad).

The IoW 1989 (IOW F2) cohort was supported by the National Heart, Lung, and Blood Institute under the award number R01 HL132321 (PI: Wilfried Karmaus).

We would like to thank all the participants of the Isle of Wight birth cohorts, the research team at David Hide Asthma & Allergy Research Centre (Isle of Wight) for collecting the data. In particular, the nurses for their help in recruiting them, Stephen Porter, Sharon Matthews, Frances Mitchell, Nikki Graham for technical support and other members of the IoW research group for valuable discussion. DNA methylation data was generated by the Oxford Genomics Centre at the Wellcome Trust Centre for Human Genetics.

**LINA**

Lifestyle and environmental factors and their Influence on Newborns Allergy risk (LINA): The LINA study is funded by the Helmholtz Centre for Environmental Research - UFZ. Parts of the LINA study were funded by the Helmholtz Impulse and Networking Fund as well as by the German Research Foundation (KFO250). EWAS analyses in the LINA study were funded by the German Cancer Research Center – DKFZ.

We cordially thank the participants of the LINA study as well as their parents, the midwives and the physicians. We are grateful to Beate Fink, Anne Hain, Michaela Loschinski, Sandra Albrecht, Melanie Bänsch and André Andrecke for their excellent technical assistance and fieldwork.

**MOBA**

MOBA: The Norwegian Mother, Father and Child Cohort Study are supported by the Norwegian Ministry of Health and Care Services and the Ministry of Education and Research, NIH/NIEHS (contract no N01-ES-75558), NIH/NINDS (grant no.1 UO1 NS 047537-01 and grant no.2 UO1 NS 047537-06A1). For this work, MoBa 1 and 2 were supported by the Intramural Research Program of the NIH, National Institute of Environmental Health Sciences (Z01-ES-49019) and the Norwegian Research Council/BIOBANK (grant no 221097). The work was partly funded by The Norwegian Research council's Centre of Excellence Scheme (grant no 262700).

The Norwegian Mother, Father and Child Cohort Study is supported by the Norwegian Ministry of Health and Care Services and the Ministry of Education and Research. We are grateful to all the participating families in Norway who take part in this on-going cohort study.

**NEST**

The NEST study was funded by NIEHS grants R21ES014947 and R01ES016772 and NIDDK grant R01DK085173. CH and DDJ received support from the Center for Human Health and the Environment grant received from the National Institute of Health Science (P30 ES025128). CH, SKM, and RLM also received support from a National Institute of Health Science grant (R24 ES028531).

We thank the parents and other caregivers of the Newborn Epigenetics STudy. We also thank the field and laboratory staff for their effort.

**NFBC1986**
NFBC1986 has received financial support from the Academy of Finland (104781, 120315, 121620, 129269, 1114194, 24300796), Center of Excellence in Complex Disease Genetics and SALVE, Oulu University Hospital, Oulu, Finland, Biocenter Oulu, Finland, University of Oulu, Finland (75617, 24002054, 2400692), Ministry of Social Affairs and Health (50459, 50691, 50842, 2749, 2465), NHLBI grant 5R01HL087679-02 through the STAMPEED program (1RL1MH083268-01), NIH/NIMH (5R01MH63706:02), ENGAGE project and grant agreement HEALTH-F4-2007-(201413), EU FP7 EurHEALTHAgeing (277849), EU FP7 EurHealth Epi-Migrant (279143), European Regional Development Fund 537/2010 (24300936) and the Medical Research Council, UK (G0500539, G0600705, G1002319, PrevMetSyn/SALVE).
Epigenetics research in the NFBC1986 received support by H2020–633595 DynaHEALTH, H2020 733206 LifeCycle, H2020-824989 EUCANCONNECT, the academy of Finland EGEA-project (285547), the Biocenter Oulu and the JPI HDHL, PREcisE project, ZonMw the Netherlands no. P75416).

We gratefully acknowledge the contributions of the participants in the Northern Finland Birth Cohort 1986. We also thank all the field workers and laboratory personnel for their efforts.

**PIAMA**

The PIAMA study is supported by The Netherlands Organization for Health Research and Development; The Netherlands Organization for Scientific Research; The Lung Foundation of the Netherlands (grant number AF 45.1.14.001 supported the methylation assays); The Netherlands Ministry of Spatial Planning, Housing, and the Environment; and The Netherlands Ministry of Health, Welfare, and Sport.

We thank all the children and their parents for their cooperation. We also thank all the field workers and laboratory personnel involved for their efforts, and Marjan Tewis for data management.

**PREDO**

Prediction and Prevention of Preeclampsia and Intrauterine Growth Restriction (PREDO): The PREDO study has received funding from the Academy of Finland, EraNet, EVO (a special state subsidy for health science research), University of Helsinki Research Funds, the Signe and Ane Gyllenberg foundation, the Emil Aaltonen Foundation, the Finnish Medical Foundation, the Jane and Aatos Erkko Foundation, the Novo Nordisk Foundation, the Päivikki and Sakari Sohlberg Foundation, the Sigrid Juselius Foundation, and the Sir Jules Thorn Charitable Trust.

The PREDO study would not have been possible without the dedicated contribution of the PREDO Study group members: A Aitokallio-Tallberg, A-M Henry, VK Hiilesmaa, T Karipohja, R Meri, S Sainio, T Saisto, S Suomalainen-König, V-M Ulander, T Vaitilo (Department of Obstetrics and Gynaecology, University of Helsinki and Helsinki University Central Hospital, Helsinki, Finland), L Keski-Nisula (Kuopio University Hospital, Kuopio Finland), E Koistinen, T Walle, R Solja, P Taipale (Northern Karelia Central Hospital, Joensuu, Finland), M Kurkinen (Päijät-Häme Central Hospital, Lahti, Finland), P Staven (Iisalmi Hospital, Iisalmi, Finland), J Uotila (Tampere University Hospital, Tampere, Finland). We also thank the PREDO cohort mothers, fathers and children for their enthusiastic participation.

**PROGRESS**

Programming Research in Obesity, Growth, Environment and Social Stressors study (PROGRESS): National Institute of Environmental Health Sciences (P30ES023515, R01ES020268, R01ES013744, R01ES014930, R01ES021357, R24ES028522; R00ES023450).

We acknowledge the American British Cowdray Medical Center for providing research facilities, which made it possible to conduct the study.

**Project Viva**

Grants from the US National Institutes of Health (R01 HD034568, UH3 OD023286, R01 HL111108, R01 NR013945).

We are grateful to families who participate in Project Viva.

**Raine**

For Raine, the DNA methylation work was supported by NHMRC grant 1059711. Collaborative analyses are supported by NHMRC 1142858. Data collection and biological specimens at the 17-year follow-up were funded by the NHMRC Program Grant ID 353514 and Project Grant 403981. RCH and TAM are supported by NHMRC Fellowship grant number 1053384 and 1136046 respectively. This work was supported by resources provided by The Pawsey Supercomputing Centre with funding from the Australian Government and the Government of Western Australia**.** This project received funding from the European Union's Horizon 2020 research and innovation programme (733206, LIFECYCLE). The authors are grateful to The Raine Study participants and their families, and The Raine Study management team for cohort co-ordination and data collection, the National Health & Medical Research Council (NHMRC) for their long-term contribution to funding the study over the last 29 years and The Telethon Kids Institute for long term support of the Study. We also acknowledge The University of Western Australia (UWA), Raine Medical Research Foundation, The Telethon Kids Institute, Women and Infants Research Foundation, Edith Cowan University, Murdoch University, the University of Notre Dame Australia, Raine Medical Research Foundation, and Curtin University for providing funding for Core Management of The Raine Study.

**STOPPA**

STOPPA: Financial support was provided by the Swedish Research Council (grant no 2018-02640) and through the Swedish Initiative for research on Microdata in the Social And Medical Sciences (SIMSAM) framework grant number 340-2013-5867, grants provided by the Stockholm County Council (ALF projects), the Swedish Heart Lung Foundation, the Swedish Asthma and Allergy Association's Research Foundation, FORTE and Stiftelsen Frimurare Barnahuset Stockholm. CA acknowledges support by the Swedish Research Council (grant number 2018-02640 and 340-2013-5867), grants provided by the Stockholm County Council (ALF projects), the Swedish Heart Lung Foundation, the Swedish Asthma and Allergy Association's Research Foundation, FORTE (grant number 2015-00289) and Stiftelsen Frimurare Barnahuset Stockholm. We acknowledge the Swedish Twin Registry for access to data. The Swedish Twin Registry is managed by Karolinska Institutet and receives funding through the Swedish Research Council under the grant no 2017-00641. We also acknowledge the Biobank at Karolinska Institutet for professional biobank service.

First, we direct our greatest appreciation to the twins and parents of the STOPPA cohort, without whose participation this study could not have been performed. We are also indebted to the STOPPA research nurses and database managers for their excellent data collection and data managing. We also want to direct our thanks to the eight paediatric allergy clinics around Sweden for their great collaboration during our visits for clinical examinations. We acknowledge the Swedish Twin Registry for access to data. The Swedish Twin Registry is managed by Karolinska Institutet and receives funding through the Swedish Research Council under the grant no 2017-00641. We also acknowledge the Biobank at Karolinska Institutet for professional biobank service.
